# COVID-19 in Connecticut institutions of higher education during the 2020-2021 academic year

**DOI:** 10.1101/2021.08.11.21261732

**Authors:** Olivia Schultes, Victoria Clarke, A. David Paltiel, Matthew Cartter, Lynn Sosa, Forrest W. Crawford

## Abstract

**Background:** During the 2020-2021 academic year, many institutions of higher education reopened to residential students while pursuing strategies to mitigate the risk of SARS-CoV-2 transmission on campus. Reopening guidance emphasized PCR or antigen testing for residential students and social distancing measures to reduce the frequency of close interpersonal contact. Connecticut colleges and universities employed a variety of approaches to reopening campuses to residential students.

**Methods:** We used data on testing, cases, and social contact in 18 residential college and university campuses in Connecticut to characterize institutional reopening strategies and COVID-19 outcomes. We compared institutions’ fall 2020 COVID-19 plans, submitted to the Connecticut Department of Public Health, and analyzed contact rates and COVID-19 outcomes throughout the academic year.

**Results:** In census block groups containing residence halls, fall student move-in resulted in a 475% (95% CI 373%-606%) increase in average contact, and spring move-in resulted in a 561% (441%-713%) increase in average contact. The relationship between test frequency and case rate per residential student was complex: institutions that tested students infrequently detected few cases but failed to blunt transmission, while institutions that tested students more frequently detected more cases and prevented further spread. In fall 2020, each additional test per student per week was associated with a reduction of 0.0014 cases per student per week (95% CI: -0.0028, -0.000012). Residential student case rates were associated with higher case rates in the town where the school was located, but it is not possible to determine whether on-campus infections were transmitted to the broader community or vice versa.

**Conclusions:** Campus outbreaks among residential students might be avoided or mitigated by frequent testing, social distancing, and mandatory vaccination. Vaccination rates among residential students and surrounding communities may determine the necessary scale of residential testing programs and social distancing measures during the 2021-2022 academic year.

## Introduction

Institutions of higher education throughout the United States ended in-person education in response to the COVID-19 pandemic in the spring of 2020^1^. Over the summer of 2020, policymakers, advocates, researchers, and university leaders developed guidelines and policies for safely reopening colleges and universities to residential students and in-person education^2-7^. Recommendations included testing (molecular, antigen, antibody), quarantine, contact tracing, social distancing, gating criteria for opening/closing, adjustments to academic terms, and masking. A major point of controversy was the importance of viral testing for residential students. The Centers for Disease Control and Prevention (CDC) published guidance that did not recommend SARS-CoV-2 viral testing upon arrival on campus^8^, which was met with objections from public health scholars^9^. Other guidelines published over the summer of 2020 did not specify a recommended frequency for testing for residential students or staff^4,6^. Even by October 2020, CDC had not explicitly recommended either entry screening or routine asymptomatic testing, though guidelines stated “Testing a random sample of asymptomatic students, faculty, and staff could increase the timeliness of outbreak detection […] CDC recommends IHEs implement both universal entry screening and expanded serial screening testing at least weekly if sufficient testing capacity is available.”^10^.

In the absence of clear recommendations from public health authorities, researchers developed mathematical models of disease transmission to predict campus COVID-19 outcomes under different testing regimes and guide university reopening plans^11-15^. These model-based analyses provided similarly varied prescriptions for safe reopening: some recommended testing only a fraction of students per week^16,17^, while others recommended at least twice weekly screening for each student^11,12^. Late in the summer of 2020, researchers warned^18^ about the risk of significant outbreaks resulting from low testing rates^14,19^ and inequalities in access to comprehensive testing^20^.

In Connecticut, where about 190,000 students are enrolled in institutions of higher education^2^, Governor Ned Lamont closed all public schools on March 17, 2020 and encouraged private schools to close and transition to remote instruction^21^. In early summer 2020, the education committee of the Reopen Connecticut Advisory Group provided detailed recommendations for fall 2020 reopening. The committee included representatives from the University of Connecticut, the Connecticut State Colleges & Universities (CSCU), and the Connecticut Conference of Independent Colleges (CCIC). The report made viral testing a central part of the reopening plan: “residential institutions must also test students upon arrival, and at appropriate intervals thereafter in accordance with prevailing public health guidance”^2^. Reopening was advised only in the event of low COVID-19 incidence both in surrounding communities and the state. Each residential institution of higher education in Connecticut was required to submit a reopening plan to the Connecticut Department of Public Health (CT DPH) addressing four topics: repopulation, monitoring, case containment, and campus shutdown^22^. Throughout summer 2020, the education committee issued additional guidance for fall 2020 in consultation with CT DPH^3^. Specific joint recommendations included a 14-day quarantine period for residential students arriving from high incidence states^7^, either testing upon arrival or documented pre-arrival testing^3^, and viral testing of 5-10% of residential students per week throughout the fall 2020 semester^3^. These recommended testing rates fell far below the levels separately recommended by transmission modeling analyses for suppression of outbreaks^11,12^. The significant anticipated costs of residential screening programs may have influenced CT DPH testing frequency recommendations^5,23,24^.

Despite conflicting recommendations and guidelines, most institutions of higher education in Connecticut reopened in fall 2020 and continued in-person residential education through spring 2021. Updated guidance for spring was released on January 8, 2021^25^, which recommended pre-arrival testing, quarantine after arrival to campus, and increased weekly residential testing through the end of February 2021. Updates on February 25^26^ and March 26^27^ extended the weekly testing recommendation through the end of the spring semester. On April 1, 2021, all Connecticut residents aged ≥16 years became eligible to receive a COVID-19 vaccine, and COVID-19 case counts began to decline statewide. As of July 12, 2021, 12 Connecticut institutions of higher education have established a policy requiring vaccination for at least some students^28,29^.

As the fall 2021 semester approaches, colleges and universities must again confront the challenges posed by SARS-CoV-2 and COVID-19. Reports of campus outbreaks and other experiences have informed institutions’ plans^30,31^, but uncertainty remains about the changing risk environment due to increased levels of immunity through natural infection and vaccination, as well as the threat posed by new variants. Though effective vaccines are available in the U.S., vaccination rates among students may not be high enough to prevent outbreaks on campus. As of July 2021, only about 600 of the approximately 4,000 colleges and universities in the U.S. have announced their intention to mandate student vaccination, leaving vaccination optional at most institutions^28^. At those schools, vaccination rates are likely to mirror national levels for young people, among whom about 50% are fully vaccinated^32^. Even the minority of schools that plan to mandate vaccination may vary widely in their degree of enforcement and willingness to grant exemptions. Most institutions of higher education will not achieve vaccination coverage rates high enough to eliminate supplementary testing and non-pharmaceutical prevention options. Assembling and evaluating the limited evidence about what did or did not work (and why) on campus last year could make a timely and important contribution to prevention portfolio design and planning for the safe return to normalcy in higher education.

Motivated by the lack of high-quality evidence regarding higher education reopening plans and outcomes, we present an in-depth study of COVID-19 reopening plans and outcomes at 18 colleges and universities in Connecticut – five public and thirteen private institutions – that reopened for at least partial instruction and had residential students during fall 2020. First, we compare institutions’ COVID-19 plans, submitted to CT DPH, with actual testing rates per student per week over the fall 2020 and spring 2021 semesters. To study the effects of institutional social distancing policies, we analyze patterns of close interpersonal contact in areas containing residential campuses throughout the year. We then consider the relationship between testing and COVID-19 case rates among residential students, as well as the relationship between campus case rates and case rates in the town where the school was located. We conclude with lessons learned and recommendations for institutions of higher education during the 2021-2022 academic year.

## Methods

We studied 18 Connecticut institutions of higher education that opened for at least partial in-person instruction and housed residential students during the fall 2020 semester. We included colleges and universities which a) offered traditional four-year bachelor’s degrees; b) belonged to either CSCU or CCIC (except University of Connecticut); and c) had at least 500 full-time enrolled undergraduate students in Fall 2020^33^. For institutions with more than one campus, we studied only the main campus housing residential students. Institutions included the main campus of the flagship state university (University of Connecticut), the four Connecticut state universities (Central, Eastern, Southern, and Western Connecticut State Universities), and 13 private universities (Albertus Magnus College, Connecticut College, Fairfield University, Mitchell College, Quinnipiac University, Sacred Heart University, Trinity College, University of Bridgeport, University of Hartford, University of New Haven, University of Saint Joseph, Wesleyan University, and Yale University). Figure 1 shows a map of the 18 universities studied in this paper.

**Figure 1:**
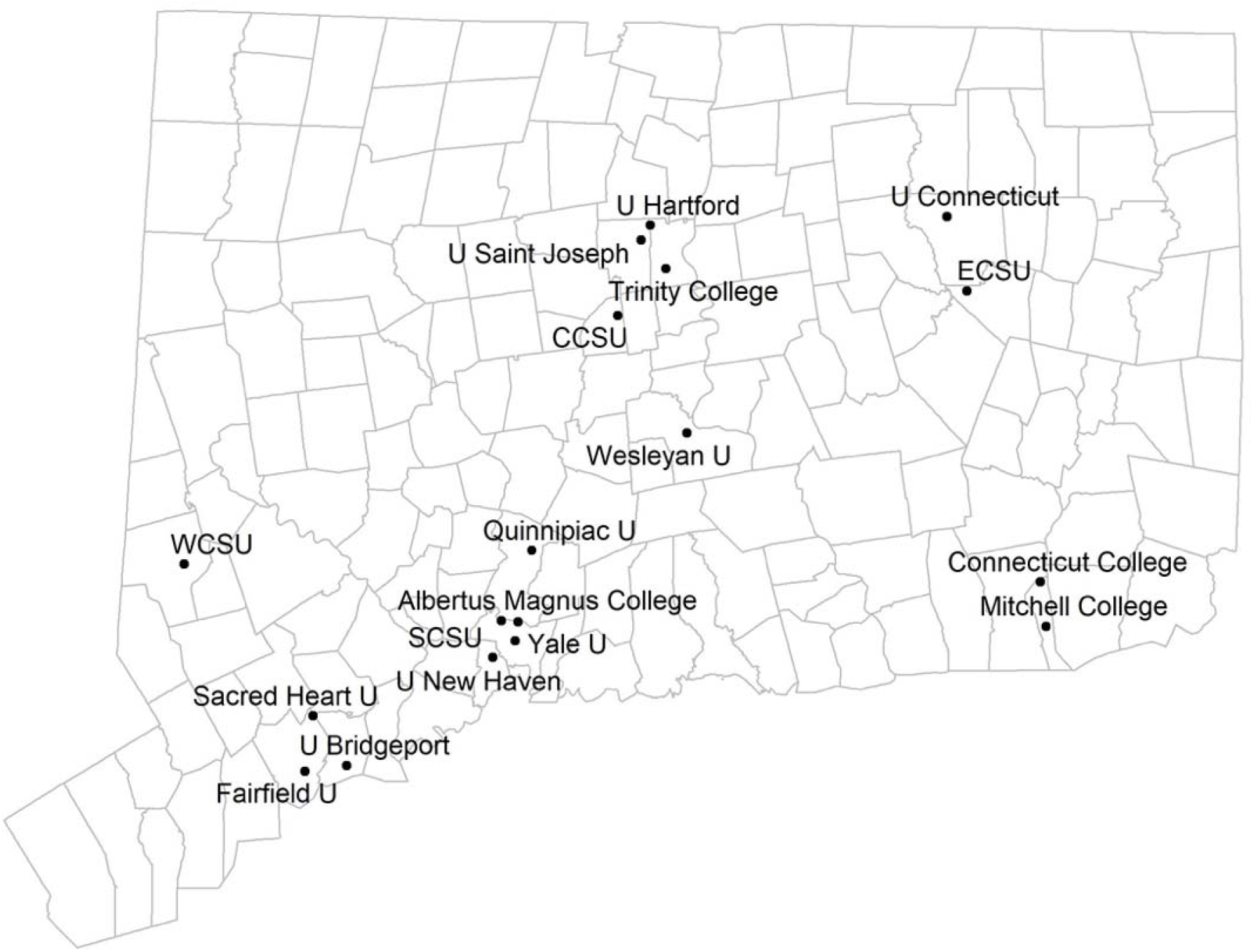
Connecticut institutions of higher education studied in this analysis.

The University of Connecticut, CSCU, and CCIC^33-36^ provided data on residential enrollment and move-in dates. Of the 18 observed colleges and universities, we obtained official reopening plans for at least one semester from all schools except Connecticut College, whose reopening plans we obtained from publicly available data. Additional data on university policies were collected from publicly available documents. Data on cost of attendance (including tuition, fees, room, and board) for residential students (2018-2019) were obtained from the 2019 Connecticut Higher Education System Data and Trends Report^37^.

Universities reported the number of COVID-19 tests conducted each week, and the number of “positive tests” recorded. In this paper, we refer to these positive tests as “cases” or “reported cases”, though it is possible that a single individual diagnosed with COVID-19 might correspond with more than one positive test. Case rates include positive tests conducted as part of university screening programs, and student-reported test results. CSCU and CCIC provided weekly data (Monday – Sunday) on student tests and cases from member institutions. Residential testing and case data from the University of Connecticut was obtained from university officials^38^ and a public dashboard^39^, which was updated daily during the fall 2020 semester and on weekdays during the spring 2021 semester. CSCU data included only polymerase chain reaction (PCR) tests with no pooling. University of Connecticut and CCIC member institutions reported both PCR and antigen tests, and University of Hartford reported pooled testing (with each individual tested reported as a single test). Campus COVID-19 testing and case data were disaggregated by residential and commuter (off-campus) students. Data on self-reported cases identified outside institutional testing programs were also included for every institution. Some data were incomplete: CSCU only collected residential testing data starting on September 20, 2020, commuter testing data starting on October 4, 2020, and self-reported cases (both residential and commuter) starting on March 14, 2021. From August 24, 2020 to October 5, 2020, Yale University reported residential testing rates more than twice the values reported in other weeks. These numbers may represent combined residential and commuter tests since reporting had just begun; data from these weeks were excluded.

We estimated the frequency of close interpersonal contact occurring on and near institutional campuses in Connecticut using mobile device data. The methodology uses location data from a cohort of mobile devices to detect likely proximity between devices within the CDC-defined 6-foot contact radius occurring over at least five minutes^40-42^. Contact counts are scaled by the estimated fraction of the total population present in the mobile device cohort, based on data from the American Community Survey^43^. The contact metric represents an estimate of the average number of contact events between distinct individuals in the population without regard to their duration or repeated contacts between the same devices. The metric excludes contact occurring in devices’ primary dwell locations (defined as locations where the device spends most of its time over the prior month) and roadways. The estimated average number of detected close contact events for each week was aggregated by census block group (CBG)^44^. For each institution, we measured contact in the CBG that most overlapped with the portion of campus containing residence halls. Campus-associated contact was defined as the contact occurring in the university-associated CBG divided by the number of residential students that semester. Not all institution-associated contact occurred on institutional campuses: contact occurring off-campus but within university-associated CBGs is included in this analysis. In figures presented below we show contact normalized by the number of residential students on campus for comparability across campuses with different residential populations; in statistical regression results we use unnormalized estimates of total contact.

From CT DPH, we obtained counts of confirmed COVID-19 cases in Connecticut towns. Data were aggregated by week and town of residential address. 2019 American Community Survey population data was downloaded from tidycensus^45^ and used to calculate weekly case rates per resident; college students living on or near campus are included in official census counts and estimates^46^. Weekly residential testing, case, and contact rates were calculated for each institution for the fall 2020 and spring 2021 semesters. We calculated these rates per residential student per week; case rates in the town where each school was located were calculated per person to ensure comparability with campus rates. We also received testing and case data for commuter students and staff/faculty from private institutions, which was used to calculate total campus cases. For contact, test, and case rates, we defined the on-campus academic period as the week of move-in through the week of move-out. In Supplemental Figure D, which explores the relationship between contact and cases, this definition was modified to end the spring semester on the last week of March, when Connecticut residents ≥16 years became eligible to receive vaccines. Weeks without data were excluded from the averages and calculation of semester-level rates. Associations between variables were estimated by least-squares linear regression. Statistical analyses were conducted using R^47^.

## Results

### University plans and policies

Table 1 shows the 18 institutions and their planned testing policies prior to the fall 2020 and spring 2021 semesters, according to public documents and plans submitted to CT DPH. Institutions brought back between 235 (Albertus Magnus College) and 4,603 (University of Connecticut) residential students in the fall, and most had fewer residential students in the spring semester. In the fall, nine institutions required that students test negative within 14 days of returning to campus, in line with recommendations from the Reopen Connecticut education committee and CT DPH^3^. Other institutions adopted stricter policies, with seven requiring tests within seven days of move-in and four testing students upon arrival. Similarly, most institutions included plans to comply with an initial 14-day quarantine for students coming from “hot spot” areas as mandated by Executive Order 7III^7,48^. Some institutions required or recommended quarantine for all students prior to (Connecticut College, Trinity College) or upon arrival (University of Connecticut, Wesleyan College, Yale University). All institutions planned on testing at least 5-10% of residential students every week as recommended by the education committee/CT DPH^3^, while Quinnipiac University stated it would test 15% per week. Connecticut College, Trinity College, Wesleyan University, and Yale University committed to testing all residential students at least once per week.

**Table 1 :**
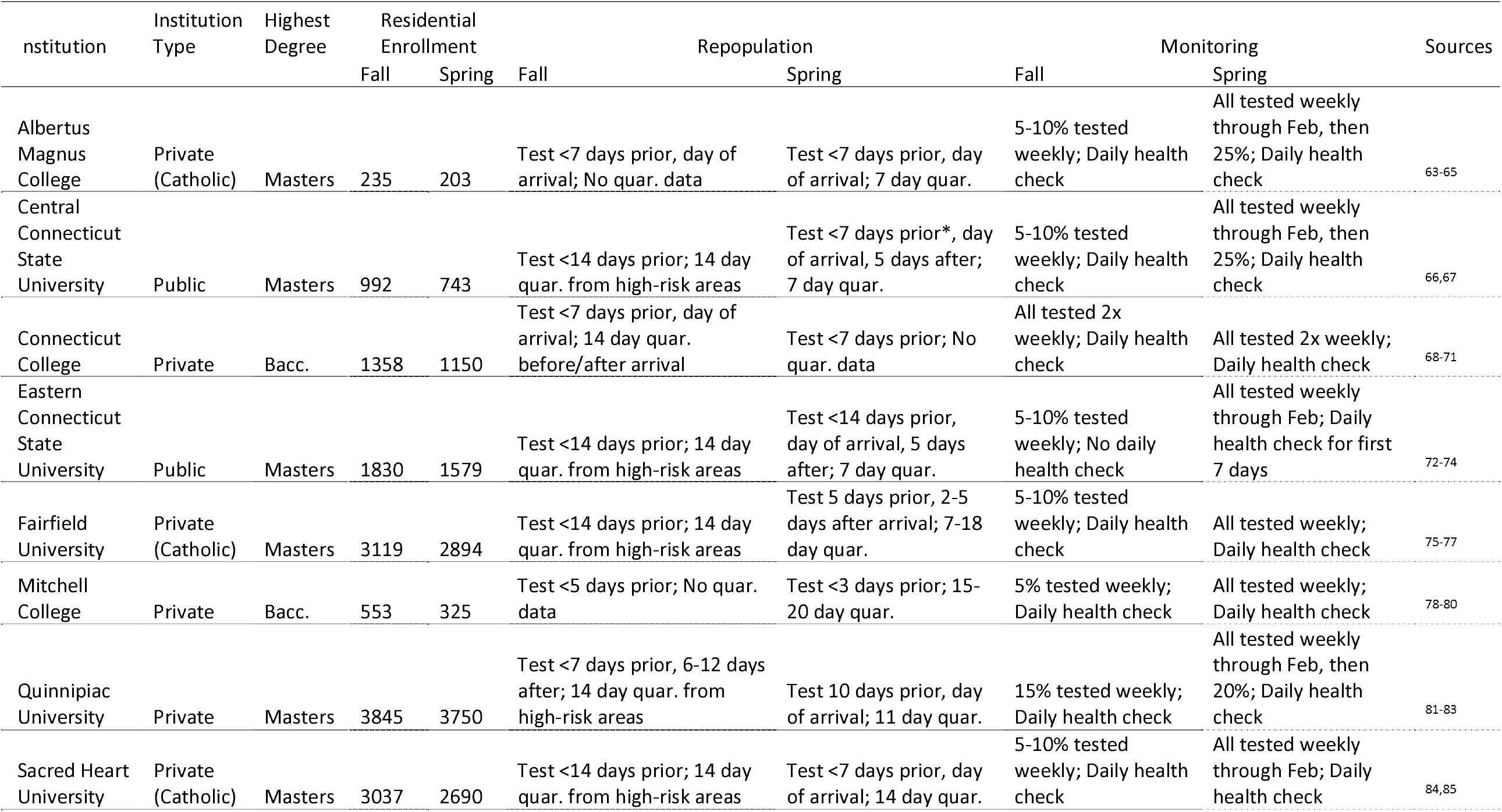
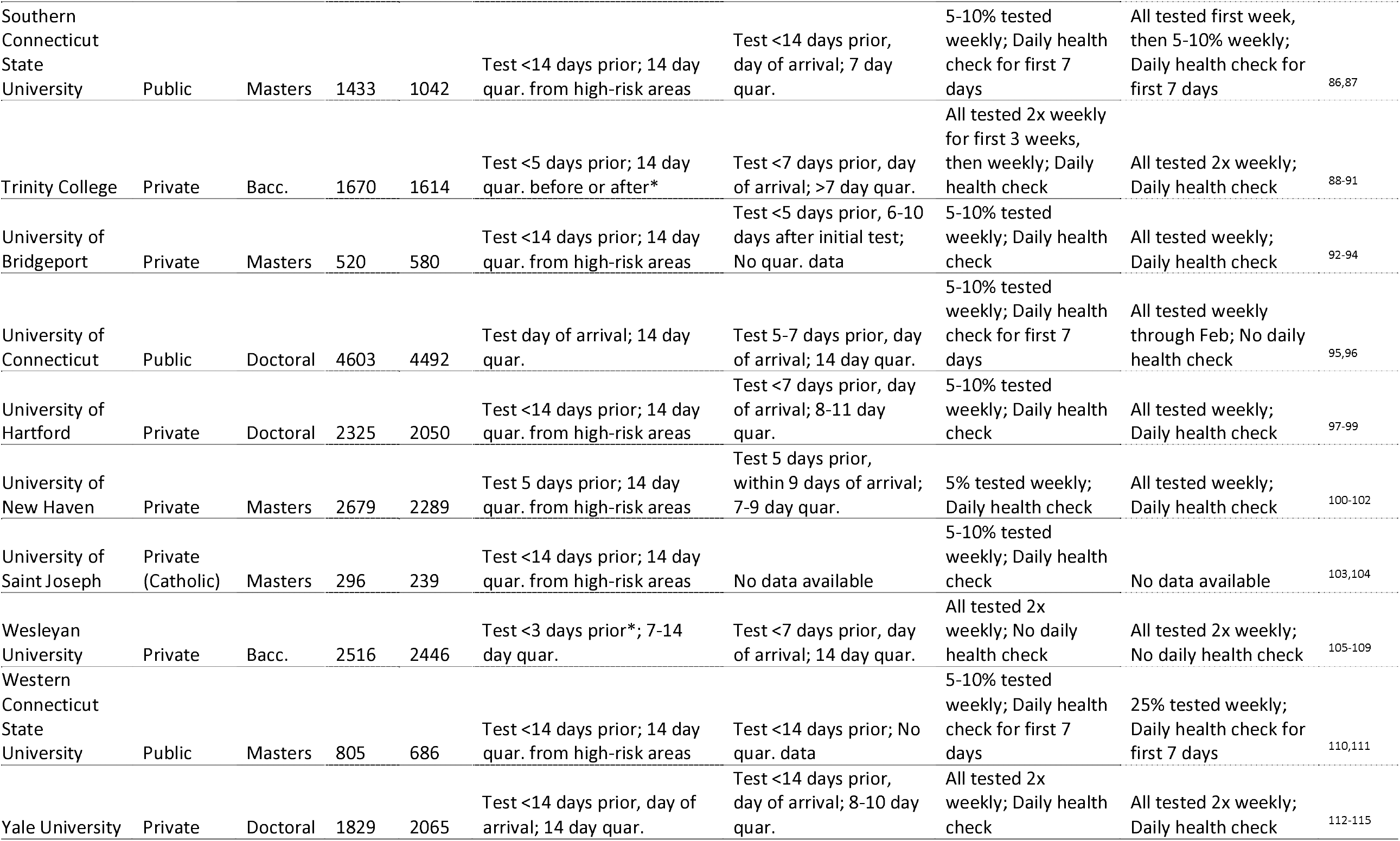
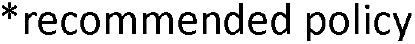
University characteristics, residential enrollment, and COVID-19 plans and policies for residential students during the 2020-2021 Academic Year. Bacc. = Baccalaureate. Quar. = quarantine

| Institution | Institution Type | Highest Degree | Residential Enrollment |  | Repopulation |  | Monitoring |  | Sources |
| --- | --- | --- | --- | --- | --- | --- | --- | --- | --- |
|  |  |  | Fall | Spring | Fall | Spring | Fall | Spring |  |
| Albertus Magnus College | Private (Catholic) | Masters | 235 | 203 | Test <7 days prior, day of arrival; No quar. data | Test <7 days prior, day of arrival; 7 day quar. | 5-10% tested weekly; Daily health check | All tested weekly through Feb, then 25%; Daily health check | 63-65 |
| Central Connecticut State University | Public | Masters | 992 | 743 | Test <14 days prior; 14 day quar. from high-risk areas | Test <7 days prior*, day of arrival, 5 days after; 7 day quar. | 5-10% tested weekly; Daily health check | All tested weekly through Feb, then 25%; Daily health check | 66,67 |
| Connecticut College | Private | Bacc. | 1358 | 1150 | Test <7 days prior, day of arrival; 14 day quar. before/after arrival | Test <7 days prior; No quar. data | All tested 2x weekly; Daily health check | All tested 2x weekly; Daily health check | 68-71 |
| Eastern Connecticut State University | Public | Masters | 1830 | 1579 | Test <14 days prior; 14 day quar. from high-risk areas | Test <14 days prior, day of arrival, 5 days after; 7 day quar. | 5-10% tested weekly; No daily health check | All tested weekly through Feb; Daily health check for first 7 days | 72-74 |
| Fairfield University | Private (Catholic) | Masters | 3119 | 2894 | Test <14 days prior; 14 day quar. from high-risk areas | Test 5 days prior, 2-5 days after arrival; 7-18 day quar. | 5-10% tested weekly; Daily health check | All tested weekly; Daily health check | 75-77 |
| Mitchell College | Private | Bacc. | 553 | 325 | Test <5 days prior; No quar. data | Test <3 days prior; 15-20 day quar. | 5% tested weekly; Daily health check | All tested weekly; Daily health check | 78-80 |
| Quinnipiac University | Private | Masters | 3845 | 3750 | Test <7 days prior, 6-12 days after; 14 day quar. from high-risk areas | Test 10 days prior, day of arrival; 11 day quar. | 15% tested weekly; Daily health check | All tested weekly through Feb, then 20%; Daily health check | 81-83 |
| Sacred Heart University | Private (Catholic) | Masters | 3037 | 2690 | Test <14 days prior; 14 day quar. from high-risk areas | Test <7 days prior, day of arrival; 14 day quar. | 5-10% tested weekly; Daily health check | All tested weekly through Feb; Daily health check | 84,85 |
| Southern Connecticut State University | Public | Masters | 1433 | 1042 | Test <14 days prior; 14 day quar. from high-risk areas | Test <14 days prior, day of arrival; 7 day quar. | 5-10% tested weekly; Daily health check for first 7 days | All tested first week, then 5-10% weekly; Daily health check for first 7 days | 86,87 |
| Trinity College | Private | Bacc. | 1670 | 1614 | Test <5 days prior; 14 day quar. before or after* | Test <7 days prior, day of arrival; >7 day quar. | All tested 2x weekly for first 3 weeks, then weekly; Daily health check | All tested 2x weekly; Daily health check | 88-91 |
| University of Bridgeport | Private | Masters | 520 | 580 | Test <14 days prior; 14 day quar. from high-risk areas | Test <5 days prior, 6-10 days after initial test; No quar. data | 5-10% tested weekly; Daily health check | All tested weekly; Daily health check | 92-94 |
| University of Connecticut | Public | Doctoral | 4603 | 4492 | Test day of arrival; 14 day quar. | Test 5-7 days prior, day of arrival; 14 day quar. | 5-10% tested weekly; Daily health check for first 7 days | All tested weekly through Feb; No daily health check | 95,96 |
| University of Hartford | Private | Doctoral | 2325 | 2050 | Test <14 days prior; 14 day quar. from high-risk areas | Test <7 days prior, day of arrival; 8-11 day quar. | 5-10% tested weekly; Daily health check | All tested weekly; Daily health check | 97-99 |
| University of New Haven | Private | Masters | 2679 | 2289 | Test 5 days prior; 14 day quar. from high-risk areas | Test 5 days prior, within 9 days of arrival; 7-9 day quar. | 5% tested weekly; Daily health check | All tested weekly; Daily health check | 100-102 |
| University of Saint Joseph | Private (Catholic) | Masters | 296 | 239 | Test <14 days prior; 14 day quar. from high-risk areas | No data available | 5-10% tested weekly; Daily health check | No data available | 103,104 |
| Wesleyan University | Private | Bacc. | 2516 | 2446 | Test <3 days prior*; 7-14 day quar. | Test <7 days prior, day of arrival; 14 day quar. | All tested 2x weekly; No daily health check | All tested 2x weekly; No daily health check | 105-109 |
| Western Connecticut State University | Public | Masters | 805 | 686 | Test <14 days prior; 14 day quar. from high-risk areas | Test <14 days prior; No quar. data | 5-10% tested weekly; Daily health check for first 7 days | 25% tested weekly; Daily health check for first 7 days | 110,111 |
| Yale University | Private | Doctoral | 1829 | 2065 | Test <14 days prior, day of arrival; 14 day quar. | Test <14 days prior, day of arrival; 8-10 day quar. | All tested 2x weekly; Daily health check | All tested 2x weekly; Daily health check | 112-115 |

\*recommended policy

CT DPH issued updated guidance for spring 2021 in early January, following several college outbreaks during the fall and increasing statewide incidence^25^. Entry testing was recommended for all residential students within 7 days of arrival to campus and again on move-in day; of the 18 institutions, 11 required a test within 5-7 days prior to move-in and 11 tested students upon arrival. The updated guidance also suggested an onboarding quarantine of 7-14 days for all residential students; 16 institutions required students to complete at least a 7-day quarantine, with five requiring 14 days. In accordance with the updated CT DPH recommendations, most institutions planned to test all residential students at least once weekly through the end of February. CT DPH weekly testing recommendations were later extended through the end of the spring semester. In addition to testing, most schools adopted a daily health check for those living or working on campus during the fall and spring semesters (13 and 12 institutions, respectively).

### Testing residential students

Figure 2 compares the planned and actual number of tests per student for each institution during fall 2020 and spring 2021, as well as the annual cost of attendance^37^. Dashed lines show the CT DPH recommended testing thresholds for each semester. During the fall 2020 semester, all institutions met or exceeded the CT DPH recommended threshold of testing at least 5% of residential students on average per week^3^. However, residential student testing rates fell short of planned and recommended thresholds during spring 2021^25-27^. Actual tests performed per residential student per week ranged from 0.18 to 2.08 in fall 2020 and from 0.38 to 1.58 in spring 2021. Connecticut College, Trinity College, Wesleyan University, and Yale University reported testing students on average more than once per week in both semesters.

**Figure 2:**
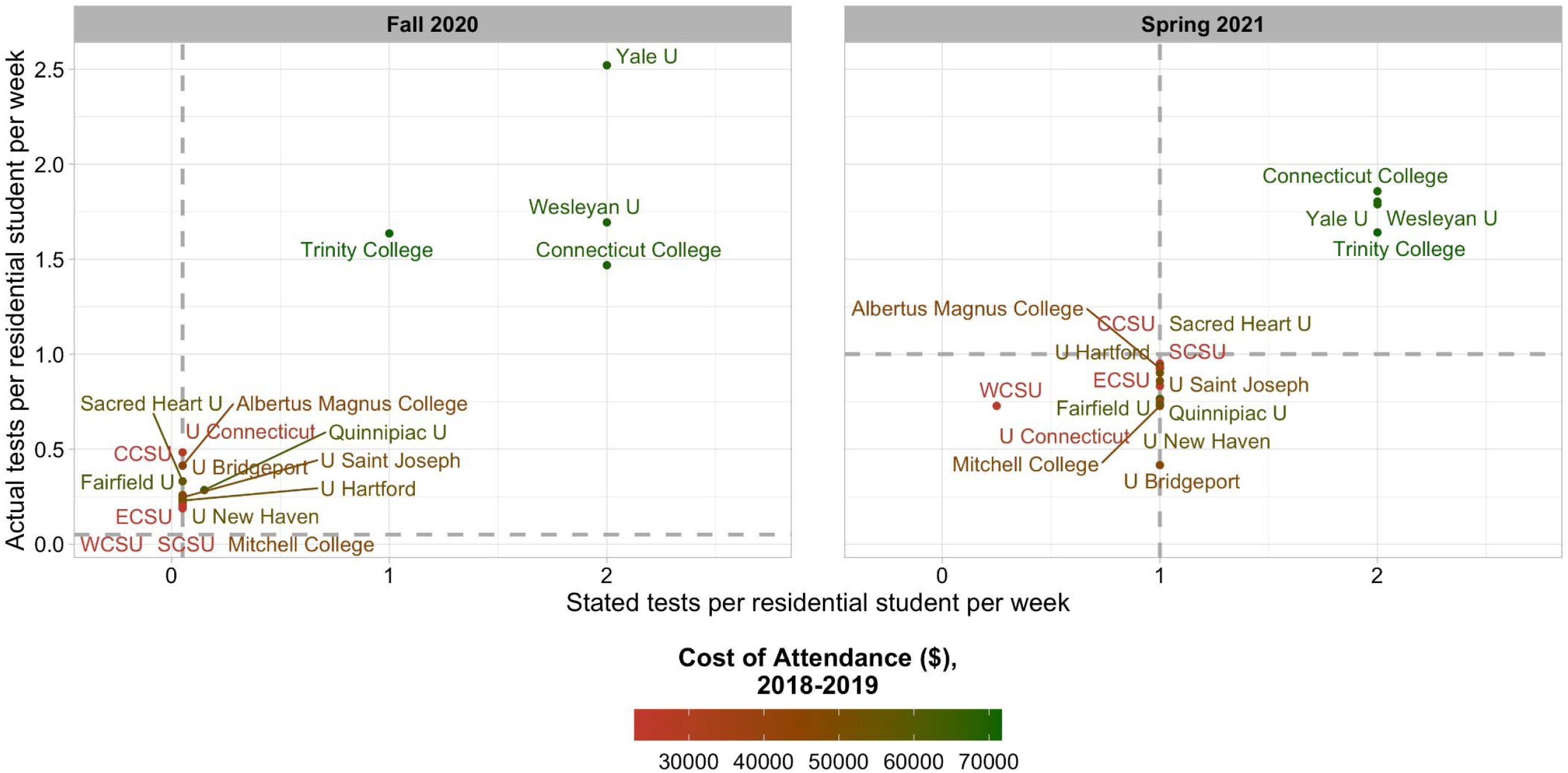
Planned and actual residential student testing rates by semester. Gray dashed lines represent CT DPH recommended testing rates for each semester.

Figure 3 shows testing rates per residential student by week during fall 2020 and spring 2021. Testing was relatively constant throughout the fall semester, although some schools conducted increased testing upon move-in (Fairfield University, University of Connecticut, University of New Haven) or move-out (Trinity College, University of Connecticut, University of Saint Joseph, Yale University), and several expanded testing during campus outbreaks (Quinnipiac University, Sacred Heart University, and University of Bridgeport). During spring 2021, testing was again mostly constant throughout the semester, with the notable exception of Albertus Magnus College which adjusted testing volume in response to several outbreaks. Albertus Magnus College, Fairfield University, Trinity College, and Yale University had increased testing during the initial quarantine period. Of the remaining institutions during spring 2021, all tested at least 75% of their students weekly except for University of Connecticut, University of New Haven, Western Connecticut State University, Mitchell College, and University of Bridgeport. Across both semesters, only Connecticut College, Trinity College, Wesleyan University, and Yale University consistently tested students roughly twice per week.

**Figure 3:**
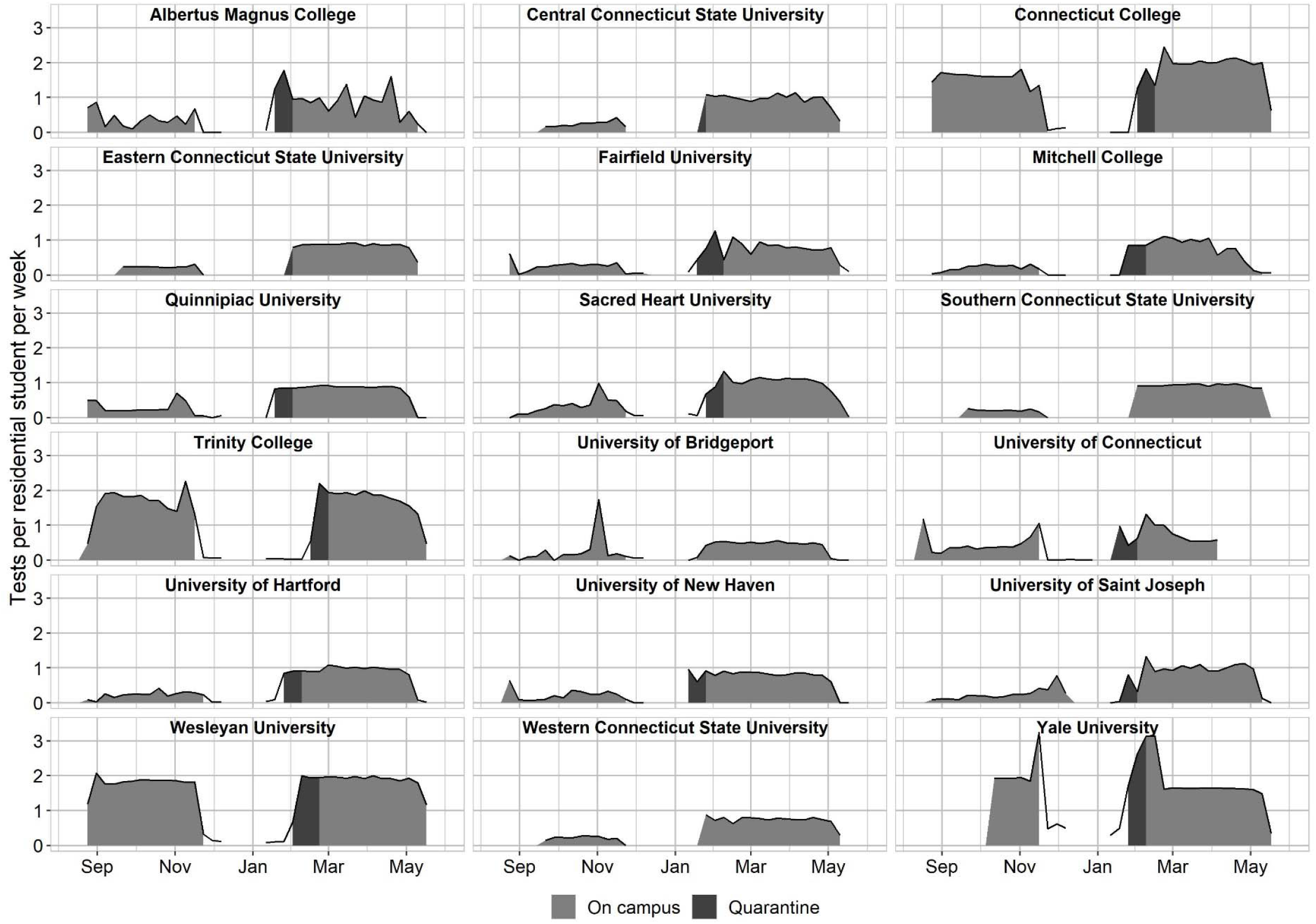
Number of COVID-19 tests per residential student per week. Shading represents weeks when students were living on campus (gray) and in quarantine (dark gray).

### Close interpersonal contact on or near campus

Close interpersonal contact increased in university-associated CBGs beginning in late August, with fall 2020 contact per residential student per week peaking at most institutions in early to mid-September. Absolute increases in contact were less substantial when students returned in the spring semester, possibly due to smaller residential enrollment or arrival lockdowns at several campuses. The four institutions testing residential students more than once per week (Connecticut College, Trinity College, Wesleyan University, and Yale University) had lower rates of contact than other schools, such as Sacred Heart University, Quinnipiac University, Fairfield University, and University of New Haven. Figure 4 shows the estimated number of close interpersonal contact events per residential student per week in university-associated census block groups. Data exclude contact occurring in primary dwell locations and roadways, and may include close contact events occurring off-campus but within university-associated CBGs.

**Figure 4:**
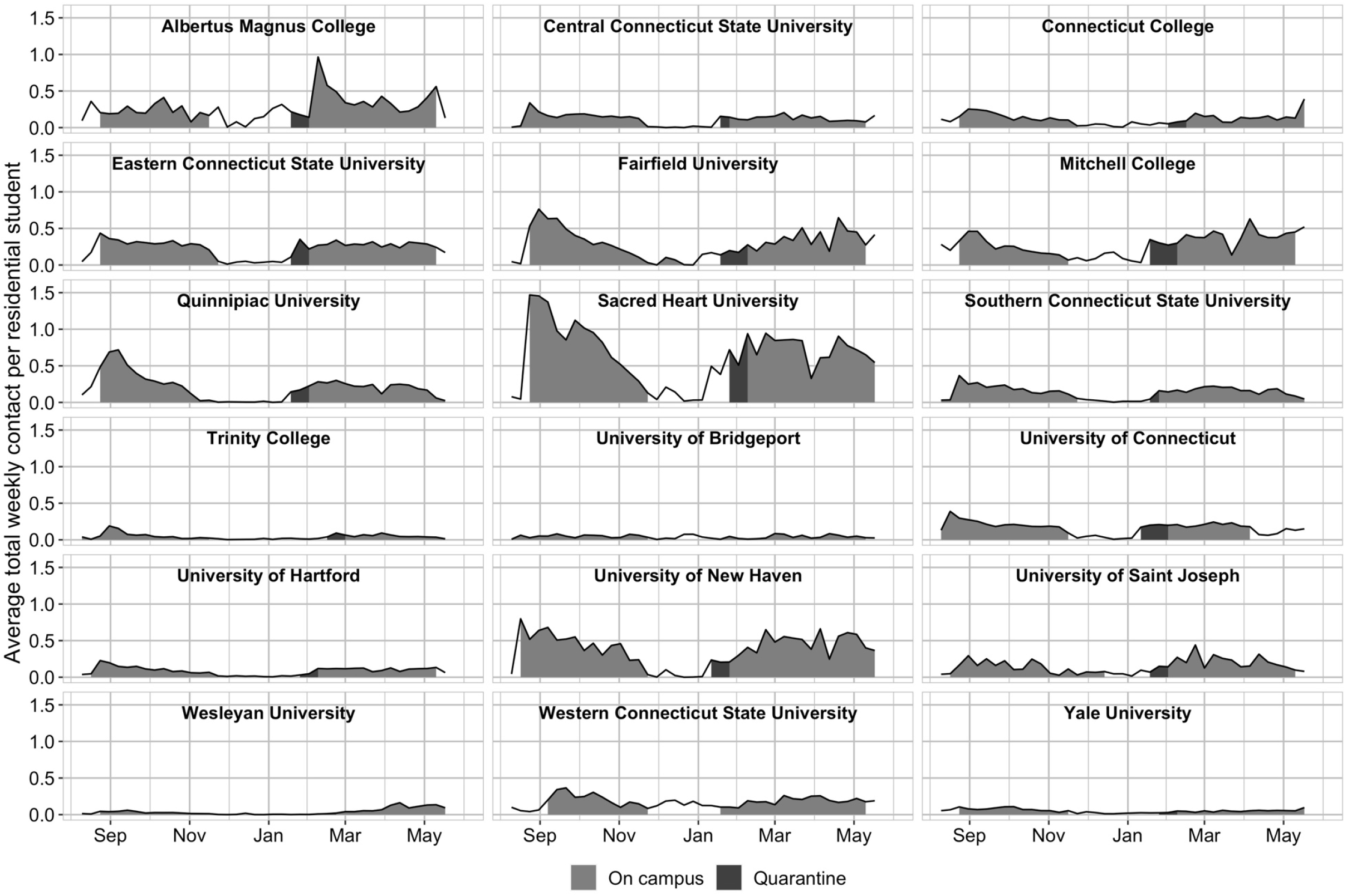
Estimated number of close interpersonal contact per residential student per week in university-associated census block groups. Shading represents weeks when students are living on campus (grey) and in initial return to campus quarantine (dark grey).

Contact rates in university-associated CBGs significantly increased during weeks when residential students were on campus: by 475% (95% CI 373%-606%) in fall 2020, and by 561% (95% CI 441%-713%) in spring 2021. Supplementary Figures A and B show contact rates as a function of time before/after move-in week. Supplementary Figure C shows estimates of the log contact rate across institutions as a function of weeks before and after the move-in date. Contact remained high for the first three weeks following campus arrival then slowly decreased throughout the rest of the semester. During spring 2021, contact increased gradually in the weeks leading up to move-in, but there was no jump in contact during move-in week or in the following weeks.

### Reported cases among residential students

Most institutions reported low residential case rates during the fall semester until early November, when most schools experienced an increase in cases. Figure 5 shows increases in the per-student case rate generally occurred before exit testing prior to move-out. Albertus Magnus College and Sacred Heart University had the highest average case rates for fall semester, while Sacred Heart University, University of Connecticut, Fairfield University, and Quinnipiac University had the highest number of absolute cases.

**Figure 5:**
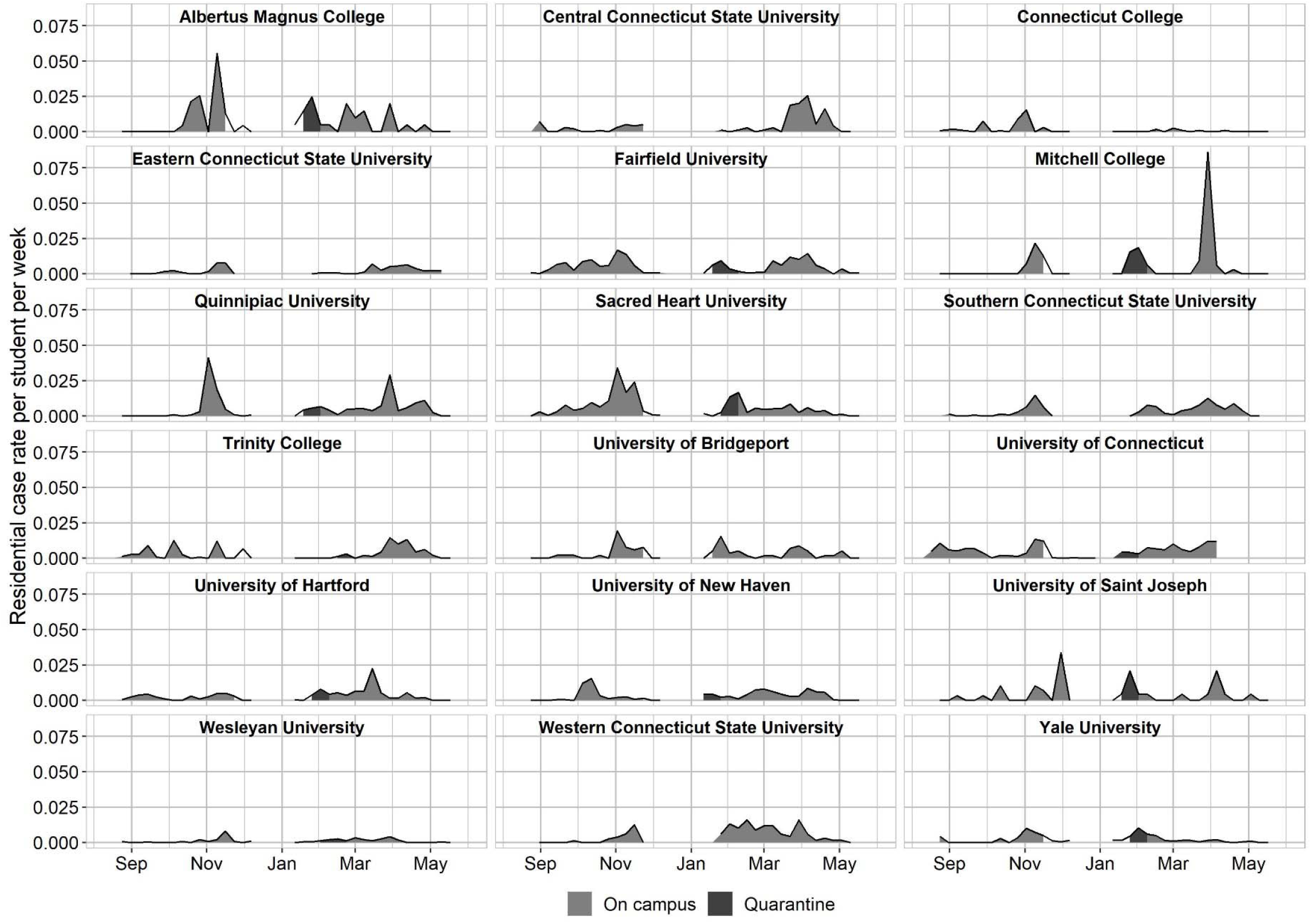
Residential student case rate per student per week by institution. Shading represents weeks when students are living on campus (gray) and in quarantine (dark gray).

Reported case patterns were more varied during spring 2021. At many institutions, initial elevated case rates decreased within a few weeks. In contrast, other institutions such as Albertus Magnus College, Central and Western Connecticut State University, and Mitchell College had continuous higher case rates or outbreaks later in the semester. These later outbreaks contributed to the highest observed spring case rates at Albertus Magnus College and Mitchell College.

Figure 6A shows that both aggregated residential case rates and testing rates were higher at most institutions during the spring semester. Institutions with higher testing rates over the entire semester had lower COVID-19 case rates in fall 2020, adjusting for the case rate in the town where the school was located. Each additional test per student per week during fall 2020 was associated with 0.0014 fewer cases per student per week (95% CI: -0.0028, -0.000012). In other words, doubling the testing rate on a hypothetical campus where 1000 residential students were tested once per week would have averted an additional 1.41 cases per week. The association between the overall semester test rate and reported case rate was not significantly different from zero during spring 2020. Figure 6B shows the relationship between residential student case rates per student per week and per-person case rates in the town where the school was located in the during fall and spring semesters. During fall 2020, the overall residential student case rate was positively associated with the town case rate, 0.0845 (95% CI: 0.0607, 0.108), while no significant relationship was observed for spring 2021. Supplementary Tables A and B show estimated regression coefficients and confidence intervals.

**Figure 6:**
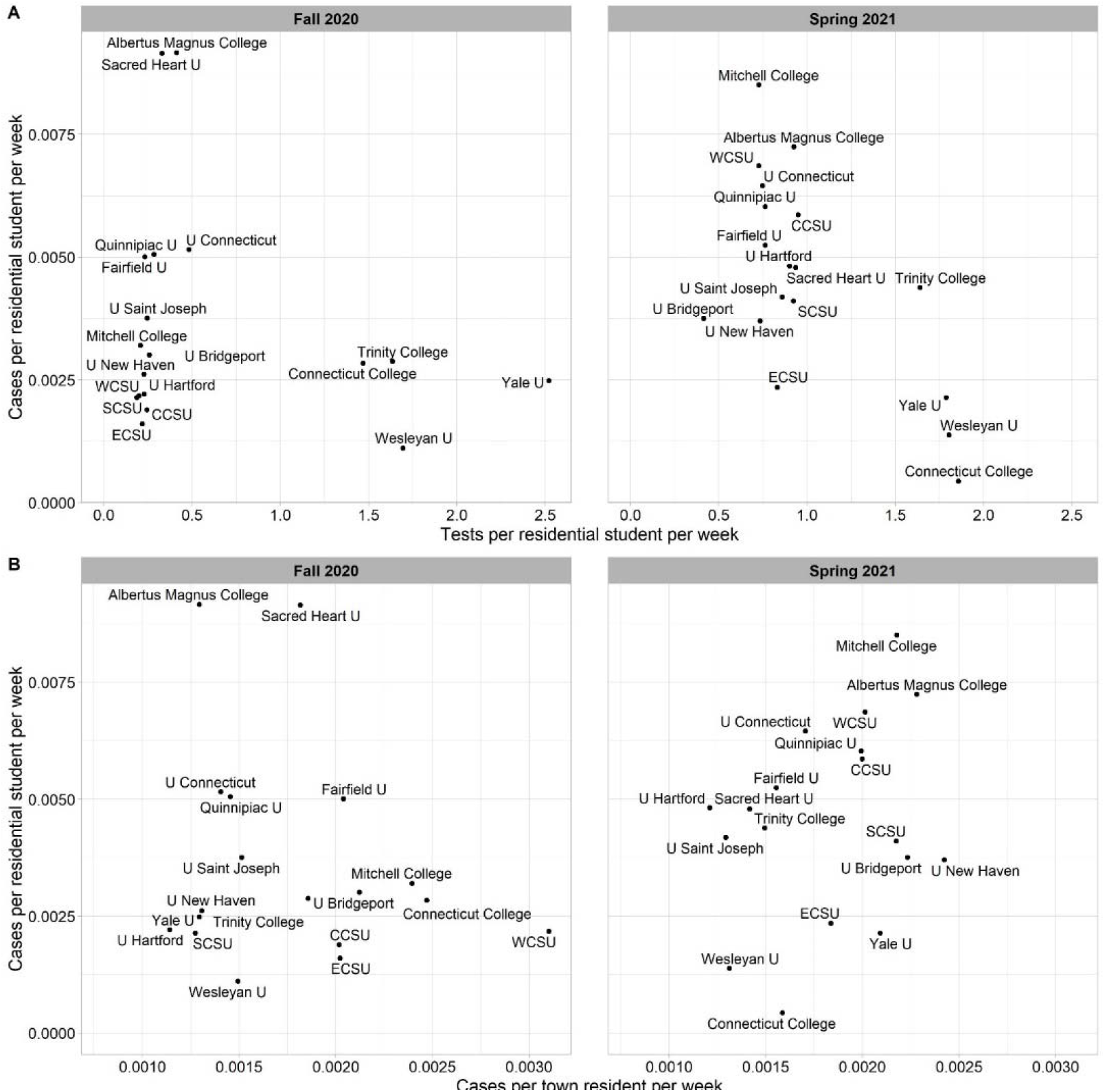
(A) Relationship between institutional test rate per residential student and case rate per residential student. (B) Relationship between case rate per residential student and case rate per person in the town where the school was located.

A weekly analysis of the relationship between test and reported case rates per residential student revealed a more complex story. Figure 7 shows that test rates below 2 times per student per week are positively associated with case rates per student, while the association for test rates above twice per week is not significantly different from zero (Supplementary Table C, Supplementary Figure D). The positive association between testing and reported cases is largest for moderate (0.5 to 1.5 tests per student per week) levels of testing, and lowest for the most infrequent (between 0 and 0.5 tests per student per week) and most frequent (more than 2 tests per student per week) testing strategies.

**Figure 7:**
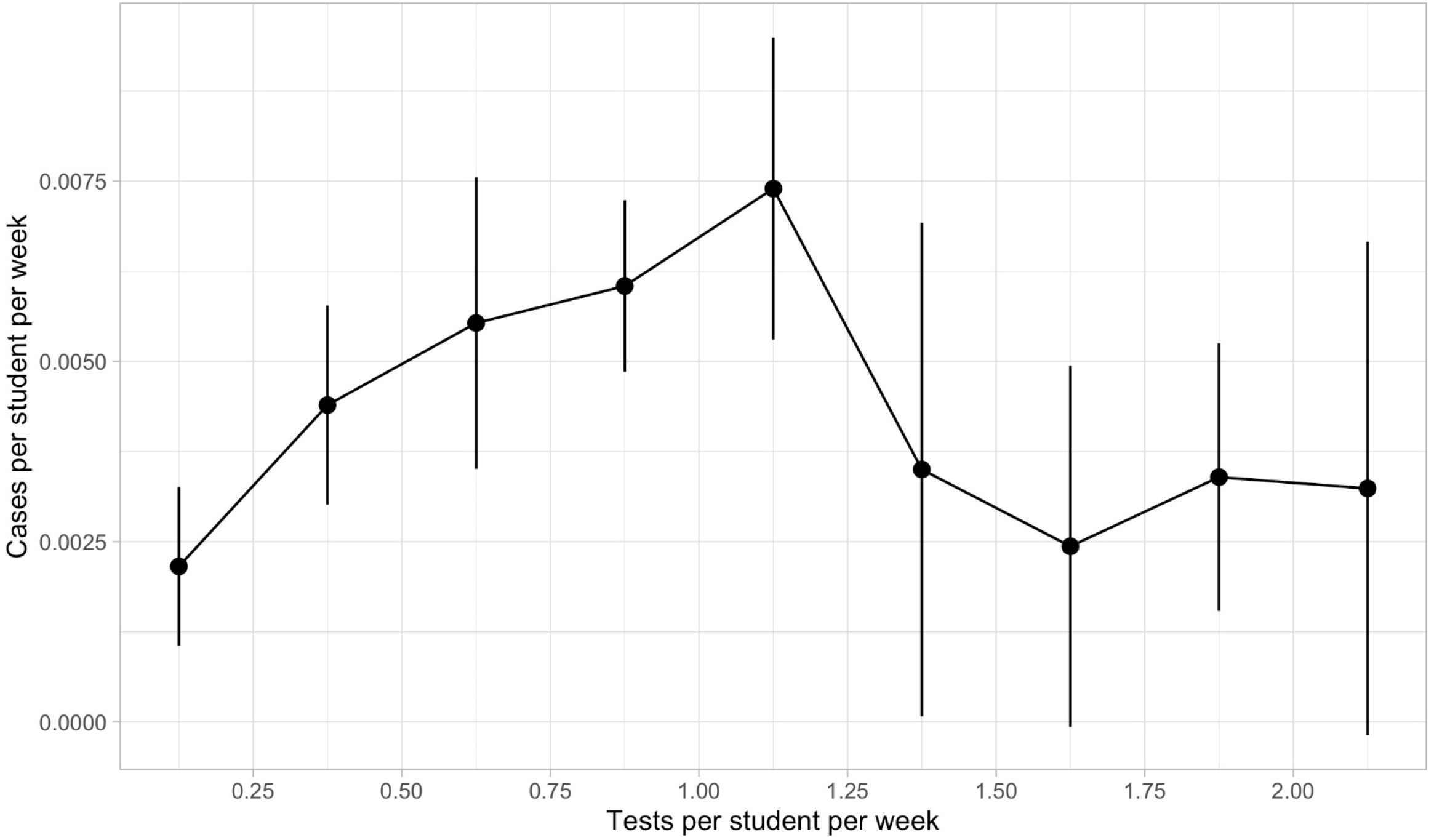
Estimates of residential cases per student per week by residential student testing rate. Estimates are represented by black dots; bars represent 95% CI.

Institutions with higher average contact per residential student throughout the semester tended to have higher reported case rates, though this relationship is stronger in the spring (Supplementary Figure E). During the spring semester the four institutions with the highest residential testing rates (Yale University, Wesleyan University, Connecticut College, and Trinity College) also had the lowest reported residential case rates.

Most residential cases were detected through institutional testing programs. Supplementary Figure F shows the relationship between university-administered tests per residential student per week and the percent of residential cases detected by the testing program. In the fall, all institutions except Albertus Magnus detected more than 75% of residential cases through their testing programs. Though testing rates increased at most institutions in the spring semester, the proportion of self-reported cases remained stable. In both semesters, the four schools testing students more than once per week detected nearly 100% of residential cases through their testing program.

We also examined total campus reported cases among residential students, commuter students, and staff/faculty in private institutions. Supplementary Figure G shows the relationship between the number of residential students and total campus cases in private universities. In both semesters, private institutions with greater numbers of residential students had more total campus cases.

## Discussion

Institutions of higher education adopted a variety of approaches to mitigating the risks of reopening residential education during a challenging 2020-2021 academic year. With the availability of effective vaccines in the U.S., the question facing higher education leaders this year is not whether there exists a risk-free approach to reopening campus; rather, the question is how to design a portfolio of vaccination, testing, contact tracing, and distancing strategies that will strike an acceptable balance between safety and the return to normalcy. Mandatory vaccination (with limited exceptions) for students and staff is the simplest approach to controlling infection on campus in the future. While many campuses have instituted mandates, vaccination remains politicized and requirements may face further legal challenges^49^. The emergence of new and more transmissible variants^50^ highlights the continued need to evaluate and implement infection control measures like testing and social distancing. Several lessons from the 2020-2021 academic year in Connecticut may provide insight that can help guide institutional plans elsewhere for safely bringing students back to campus during fall 2021.

Conflicting guidance related to the frequency of campus testing may have contributed to low testing rates at some institutions, especially during fall 2020. Lack of clarity from CDC guidelines^8^ may have led to uncertainty among policymakers and educational leaders about the importance of testing. In Connecticut, recommendations from the Reopen CT education committee^3^ and CT DPH^25-27^ set testing thresholds far below those recommended by transmission modeling studies^11,12^ to prevent campus outbreaks. These thresholds were also set during early summer 2020 when testing capacity was limited; on June 1, 2020, 9,252 tests were conducted in Connecticut, compared with 22,198 on June 1, 2021^51^. The Reopen CT education committee acknowledged that screening programs could “impose a considerable burden on institutions, especially tuition-dependent private colleges and universities, that are already coping with substantial incremental costs and revenue shortfalls arising from the pandemic”^2^. Institutions that met the minimum recommendation during fall 2020 generally experienced higher case rates than those that tested at least once per week. While frequent testing in residential institutions may be costly, testing differences across institutions were only partly explained by residential tuition and fees. Several private universities, including Quinnipiac University and Fairfield University, have high tuition but tested residential students near the CT DPH recommended minimum rate.

One of the key findings of this study is the complex relationship between residential student testing and recorded residential COVID-19 cases. Testing rates below 0.5 per student per week likely detected few asymptomatic infections; as a result, asymptomatic infectious students were not identified and isolated, so prevalent infections could spread unabated. Testing rates above 1.5 per student per week (Yale University, Wesleyan University, Connecticut College, and Trinity College in spring 2021) likely detected most infections (including asymptomatic infection) among residential students, resulting in isolation of infected individuals and suppression of further transmission. Both scenarios – low and high rates of per-student testing – resulted in low reported case rates. In contrast, moderate testing rates between 0.5 and 1.5 tests per student per week resulted in the highest reported case counts. One explanation for this finding is that moderate testing rates are frequent enough to detect many prevalent infections, but not frequent enough to stop most forward transmission and outbreaks. Our finding of similar case rates under both low and high testing rates does not mean that testing is ineffective in suppressing infection. Rather, low testing misses most infections, failing to stop transmission, while high testing captures most infections, slowing forward transmission.

Evidence regarding transmission between university campuses and the broader community suggests the possibility that infections on campus caused an increase in community incidence^52-55^. We observed a positive association between residential student case rates and case rates in the town where the school was located during fall 2021. However, it is not possible to determine whether on-campus infections were transmitted to the broader community or vice versa. While some institutions insulated their residential populations from the surrounding area via quarantine and lockdown, especially following students’ initial arrival on campus, a substantial association in case rates persists over the fall semester, indicating the possibility of cross-transmission between residential students and community members. We conclude that it may not be possible to prevent infection from spreading between campuses and surrounding communities, even under near-quarantine conditions. Institutions must be prepared to detect on-campus outbreaks arising from off-campus contact, and to prevent the spread of on-campus outbreaks to the community.

Our analysis of contact rates showed dramatic disparities in compliance with social distancing guidelines within and around campuses. Fairfield University and Sacred Heart University showed the highest contact rates in fall 2020. Both institutions experienced outbreaks on campus within weeks of residential student move-in. Across all institutions, contact rose over 400% following move-in. Close interpersonal contact is a necessary but not sufficient condition for COVID-19 transmission to occur. Both prevalent infections and high contact rates are needed to spark an outbreak. Institutions can control prevalent infection through frequent asymptomatic testing and rapid isolation of identified cases, while control of contact rates is achieved through social distancing guidelines and limits on gatherings.

Connecticut residents aged ≥16 years became eligible for vaccination on April 1, 2021, but we do not have information on vaccination rates on Connecticut campuses during the time frame studied in this paper. It is likely that rising vaccination rates on campus mitigated transmission during the remaining weeks of spring 2021, but vaccination rates among residential students during fall 2021 may not be sufficient to prevent outbreaks on campus^28,32^. CDC recommendations for institutions of higher education for fall 2021 appropriately emphasize the importance of maximizing vaccination coverage^56^. For “fully vaccinated” campuses, current CDC guidelines permit a return to pre-COVID normalcy. For campuses with a mixed population of vaccinated and unvaccinated students, CDC recommend a “layered” approach but do not offer evidence to guide the layering or to understand the tradeoffs inherent in any layering design. New CDC guidelines on testing at institutions of higher education recommend universal entry screening and asymptomatic screening, once or twice per week based on risk in the surrounding community, with priority shifting to unvaccinated persons as vaccine coverage increases^57^. Guidelines published by the American College Health Association recommend weekly testing for unvaccinated students but do not make a clear general recommendation on testing: “how frequently to test asymptomatic individuals will vary depending on the level of immunity achieved on campus either through vaccination or recovery from natural disease”^58^. The Massachusetts higher education testing group recognizes possible differences in institutions’ approaches but recommends weekly testing of all students, regardless of vaccination status^59^.

This study is subject to several limitations. First, we have focused on diagnosed COVID-19 cases recorded and reported by universities. However, cases may be a poor proxy for infections in institutions where testing rates per student were low or where asymptomatic testing was not commonplace or infrequent. It is likely that institutions with frequent asymptomatic testing of students detected essentially all infections, asymptomatic and symptomatic. At those institutions, reported case counts might be a reliable proxy for incidence of infection. In contrast, institutions that relied solely on symptomatic case reporting (and testing capacity in surrounding towns, which was dominated by self-initiated testing on demand) likely missed asymptomatic infections. For this reason, case incidence rates might not be comparable across campuses that adopted different testing strategies. Second, we used measures of close interpersonal contact as a proxy for COVID-19 transmission risk. We aggregated contact in university-associated CBGs, which may include contact occurring off-campus among non-students. Contact rates are computed from close-proximity events within a sample of mobile devices providing geo-location information^40^. However, this sample might not be representative of residential students in university-associated CBGs, or might fail to detect contact events that result in COVID-19 transmission. Contact events occurring within device primary dwell locations were excluded, so contact between students in the same residence hall may not be represented. Third, transmission on campus is influenced by a broad range of factors, including behavior of faculty and staff, as well as non-residential students on campus. Testing data for faculty, staff, and non-residential students at public universities were not routinely reported to CT DPH. These populations also might have been more likely than residential students to seek testing outside of university testing programs, given that public, free testing options were widely available in Connecticut during the 2020-2021 academic year^60^. We also observed significant variation in the rate of self-reported tests (obtained by students at non-university testing sites), possibly indicating different reporting policies for this type of test across institutions. This analysis was restricted to viral testing and therefore did not consider the impact of wastewater surveillance conducted at the University of Connecticut, which could complement future viral testing efforts in university outbreak management plans^61,62^. Finally, all Connecticut residents aged ≥16 years became eligible to receive a vaccine on April 1, 2021. This age group includes most students at colleges and universities. Statewide COVID-19 cases dropped dramatically through Spring 2021, and it is possible that during late spring, changes in broader transmission dynamics drove reductions in campus case counts more than individual institutional testing strategies or policies.

## Data Availability

COVID-19 testing and case data are publicly available from CT Department of Public Health:
https://data.ct.gov/Health-and-Human-Services/COVID-19-Tests-Cases-and-Deaths-By-Town-/28fr-iqnx
Aggregated weekly campus testing data were provided to us by the Connecticut Department of Public Health, the University of Connecticut, the Connecticut State Colleges & Universities (CSCU), and the Connecticut Conference of Independent Colleges (CCIC). These data
are not publicly available.

## Acknowledgements

We are grateful to Jacqueline Barbieri, Maciej Boni, Jessica Brockmeyer, Jared Campbell, Samantha Dean, Alexandra Edmundson, Edward H. Kaplan, Albert I. Ko, Suzanne Onorato, Alice Pritchard, Maura Provencher, William Shea, Tom Valleau, and Jennifer Widness. We thank the University of Connecticut, the Connecticut State Colleges & Universities, and the Connecticut Conference of Independent Colleges for providing data on residential enrollment, reopening plans, testing, and cases.

## Supplementary Figures

**Supp Figure A:**
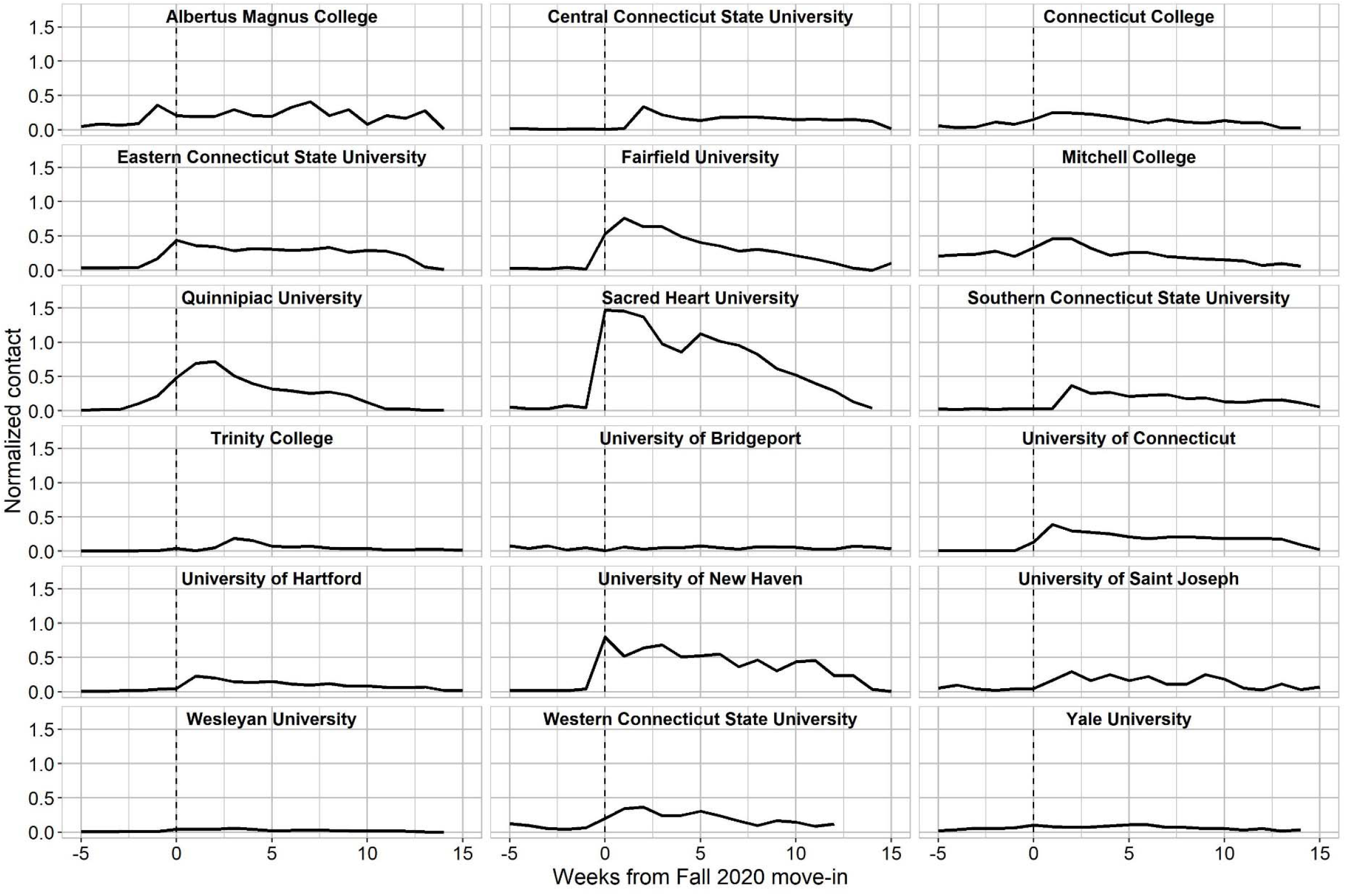
Estimated frequency of close interpersonal contact per residential student in university-associated census block groups from move-in week (week 0), fall 2020.

**Supp Figure B:**
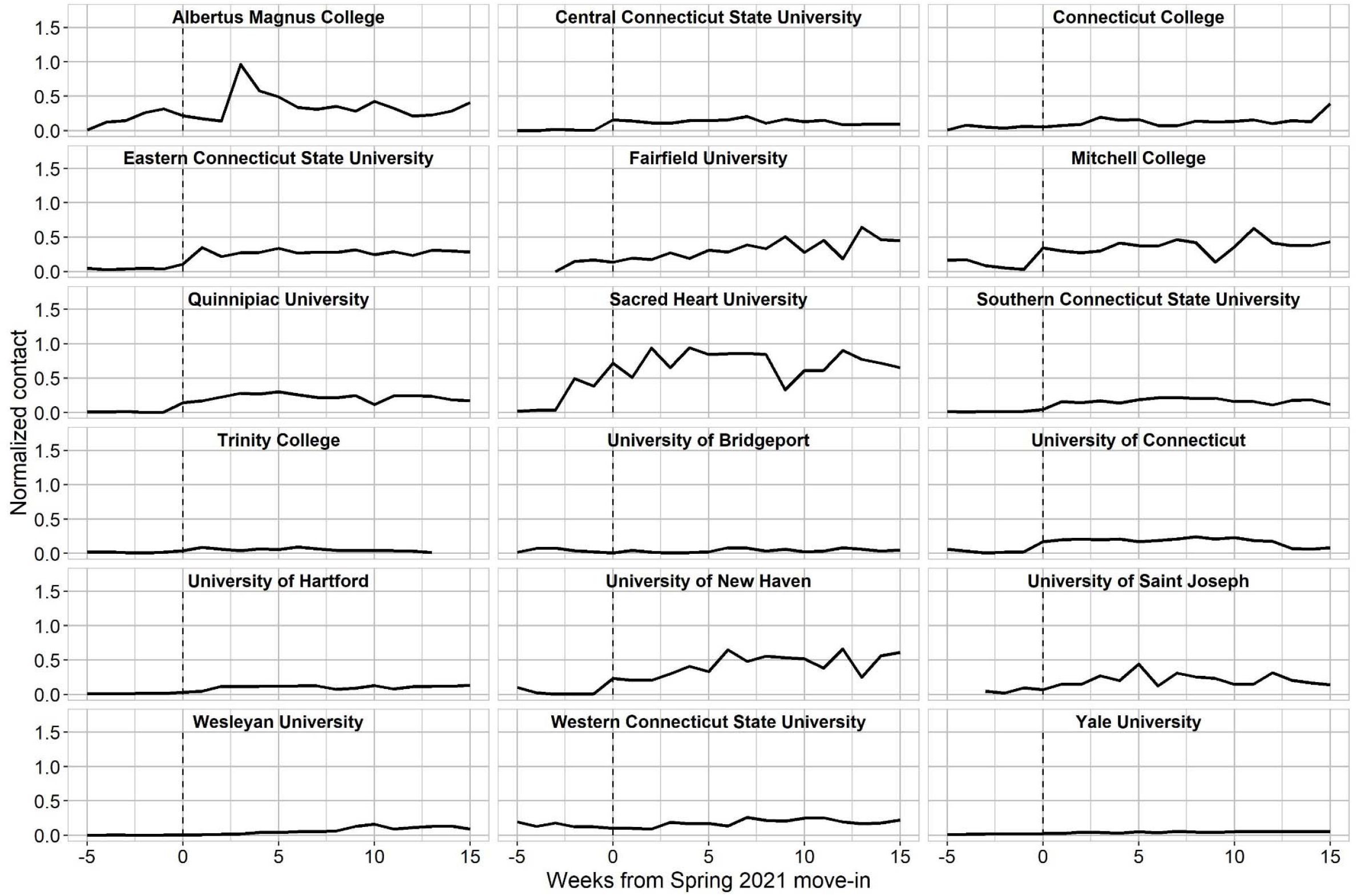
Estimated frequency of close interpersonal per residential student in university-associated census block groups from move-in week (week 0), spring 2021.

**Supp Figure C.**
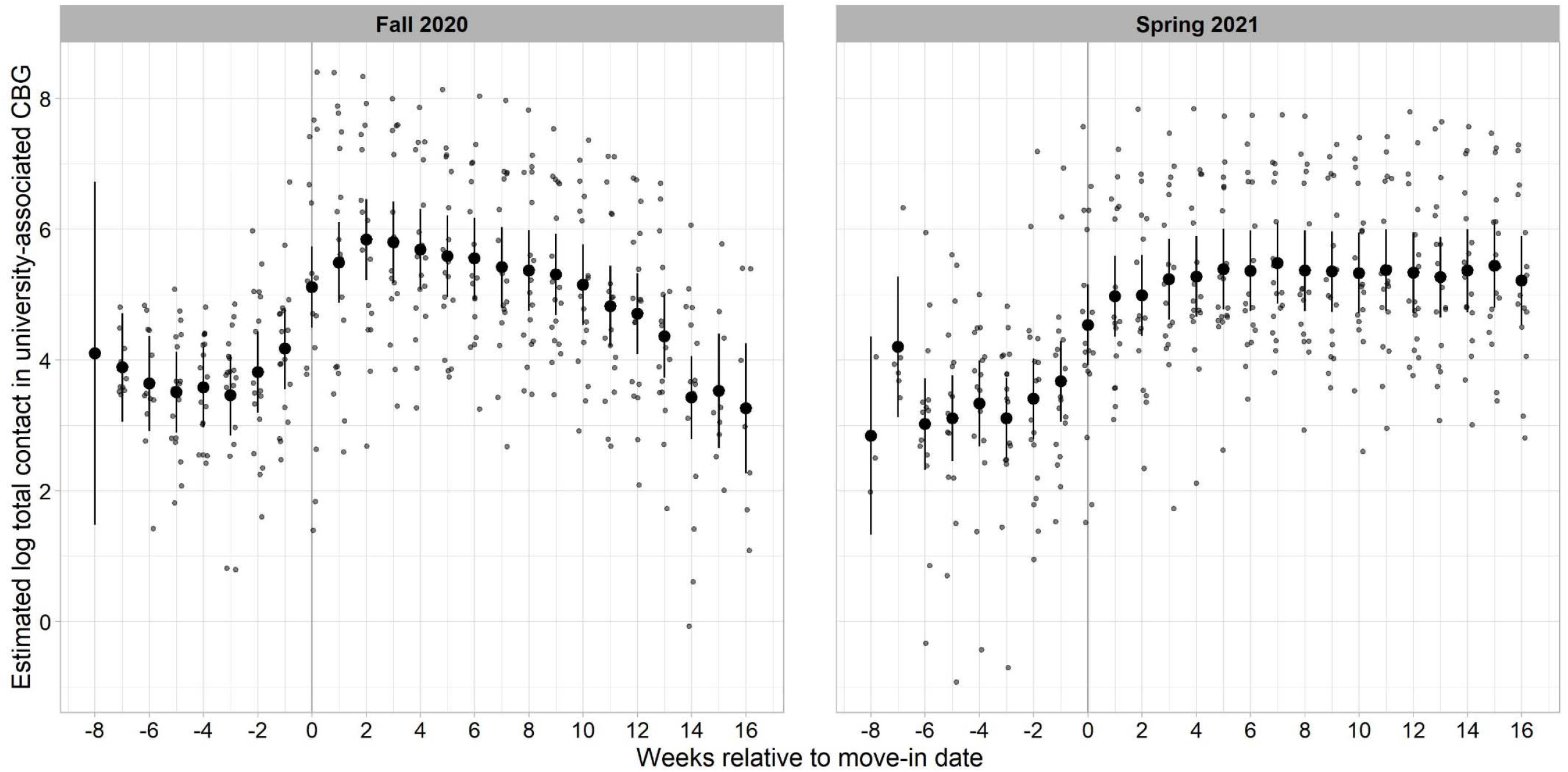
Estimated contact per residential student per week since move-in across institutions.

**Supplementary Figure D.**
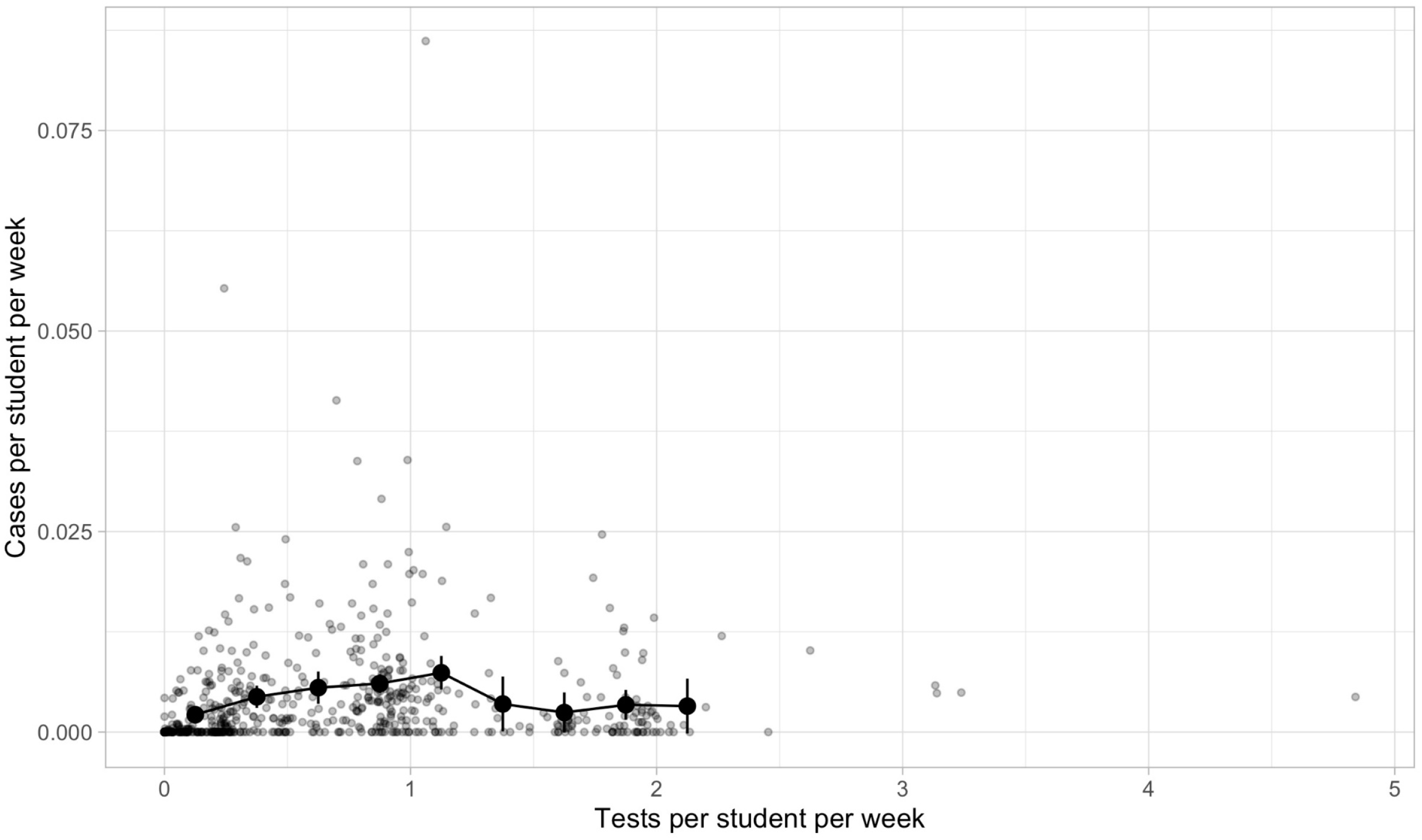
Estimates of residential cases per student per week by residential student testing rate, with weekly institution data overlaid (gray dots). Estimates are represented by black dots; bars represent 95% CI.

**Supp Figure E.**
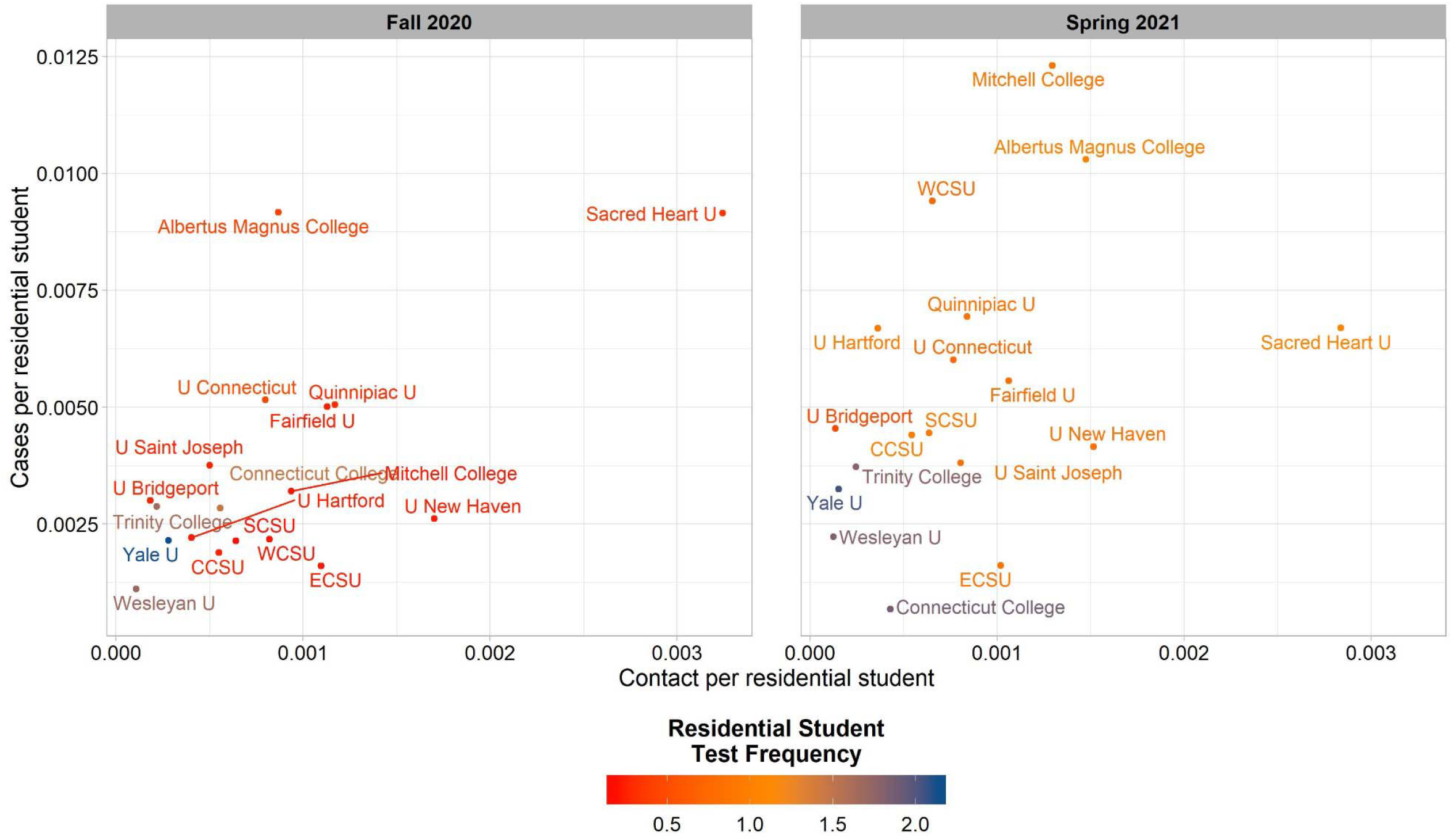
Cases per residential student as a function of contact per residential student. Fall 2020 includes data from full weeks between move-in and move-out, while spring 2021 includes data from move-in through the last week of March when college-age individuals became eligible to receive the COVID-19 vaccine in Connecticut.

**Supp Figure F:**
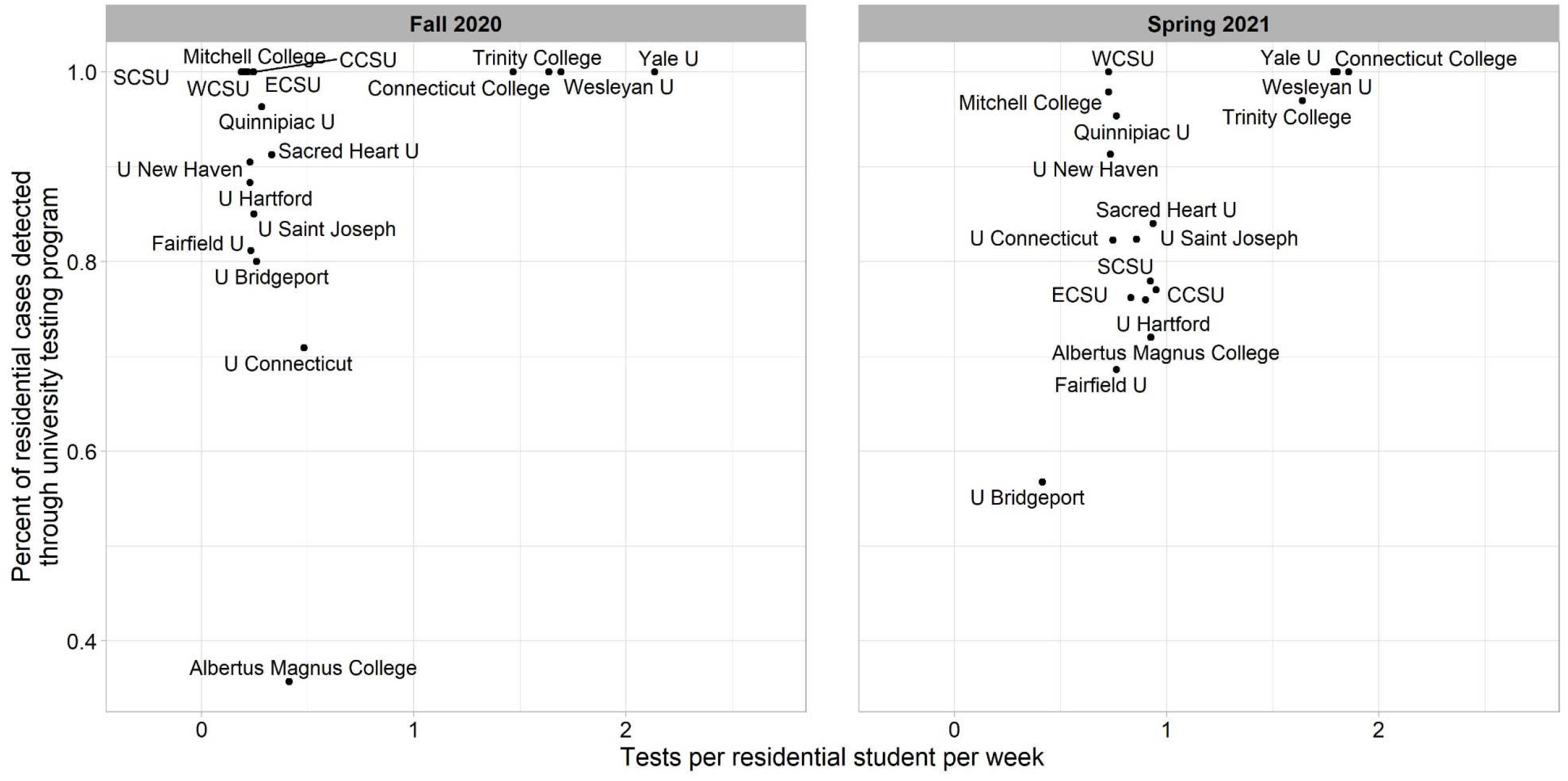
Residential student testing rate and percent of residential cases detected through university testing program, by semester. Semesters include data from full weeks between move-in and move-out.

**Supp Figure G:**
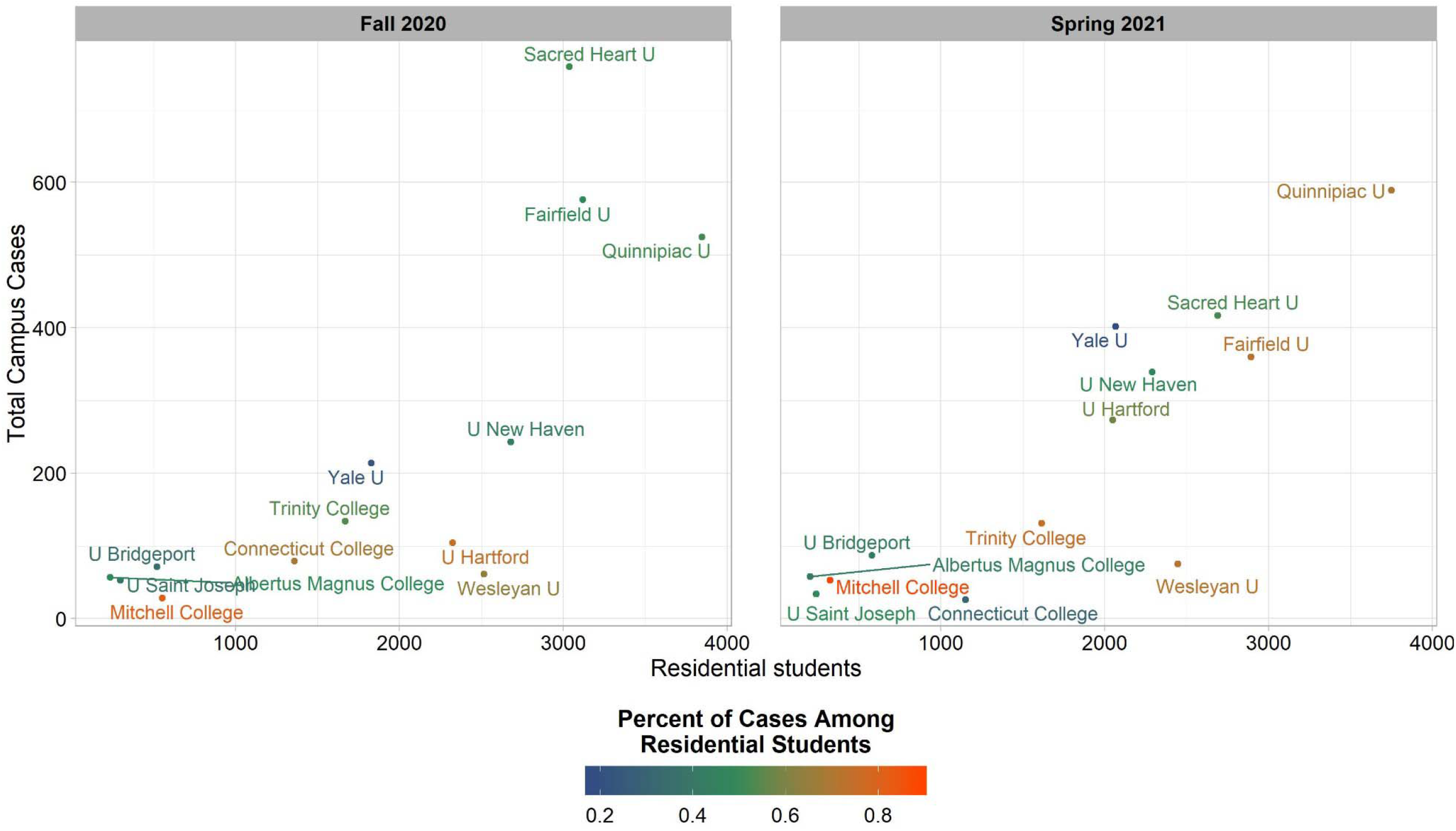
Residential students and total campus cases among private universities, by semester. Total campus cases include reported cases among residential students, commuter students, and staff/faculty. Semesters include full weeks between move-in and move-out.

**Supp Table A.**
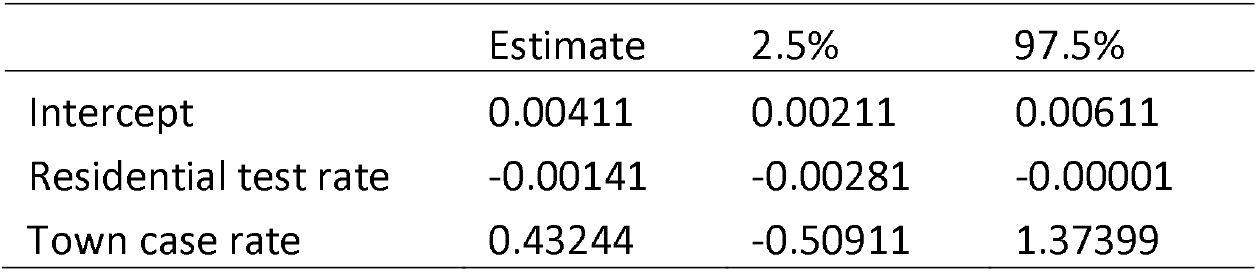
Linear regression estimates and 95% confidence intervals describing the negative association between residential test rate (per student per week) and residential case rate (per student per week), adjusted for town case rate per person.

**Supp Table B.** Linear regression estimates and 95% confidence intervals describing the association between residential case rate (per student per week) and town case rate (per person per week). The association is estimated to be positive during fall 2020, and not significantly different from zero during spring 2021.

|  | Estimate | 2.5% | 97.5% |
| --- | --- | --- | --- |
| Intercept | 0.00182 | 0.00169 | 0.00195 |
| Fall 2020 residential case rate | 0.08446 | 0.06065 | 0.10827 |
| Spring 2021 residential case rate | 0.01871 | -0.00228 | 0.03970 |

**Supp Table C:** Association between tests per student per week and cases per student per week, by test frequency.

| Tests per student per week | Estimate | 2.50% | 97.50% |
| --- | --- | --- | --- |
| 0 – 0.25 | 0.00216 | 0.00106 | 0.00326 |
| 0.25 – 0.5 | 0.00439 | 0.00301 | 0.00578 |
| 0.5 – 0.75 | 0.00553 | 0.00351 | 0.00755 |
| 0.75 – 1 | 0.00605 | 0.00485 | 0.00724 |
| 1 – 1.25 | 0.00740 | 0.00530 | 0.00949 |
| 1.25 – 1.5 | 0.00350 | 0.00008 | 0.00692 |
| 1.5 – 1.75 | 0.00244 | -0.00007 | 0.00494 |
| 1.75 – 2 | 0.00340 | 0.00154 | 0.00525 |
| >2 | 0.00324 | -0.00018 | 0.00666 |

## References

1. Hess AJ. How coronavirus dramatically changed college for over 14 million students. CNBC; 2020.

2. Levin R, Lorimer LK, Kaplan S, et al. Report of the Higher Education Subcommittee Reopen Connecticut. The Office of Governor Ned Lamont; 2020.

3. Ojakian M. Update #6 to the Higher Education Report: Recommendations for Reopening Undergraduate Colleges and Universities. Interim Guidance for Testing Higher Education Residential Students and Residence Hall Directors: Higher Education Reopen Connecticut Subcommittee; 2020.

4. National Governors Association. Memorandum: reopening institutions of higher education. National Governors Association; 2020.

5. Murakami K. Whom to test and how to pay for it. Inside Higher Ed.; 2020.

6. American College Health Association. Considerations for Reopening Institutions of Higher Education in the COVID-19 Era. ACHA; 2020:1–27.

7. Ojakian M. Update #10 to the Higher Education Report: Recommendations for Reopening Undergraduate Colleges and Universities. Guidance Related to Students from Affected States: Higher Education Reopen Connecticut Subcommittee; 2020.

8. Centers for Disease Control. Interim guidance for administrators of US institutions of higher education. 2020.

9. Bergstrom CT. The CDC Is Wrong: Testing is essential for colleges to reopen safely. The Chronicle of Higher Education. 2020.

10. Redden E. CDC Issues New Testing Guidance for Colleges. Inside Higher Ed.; 2020.

11. Chang JT, Crawford FW, Kaplan EH. Repeat SARS-CoV-2 testing models for residential college populations. Health Care Management Science. 2021;24(2):305–318.

12. Paltiel AD, Zheng A, Walensky RP. Assessment of SARS-CoV-2 Screening Strategies to Permit the Safe Reopening of College Campuses in the United States. JAMA Netw Open. 2020;3(7):e2016818.

13. Losina E, Leifer V, Millham L, et al. College campuses and COVID-19 mitigation: clinical and economic value. Annals of internal medicine. 2021;174(4):472–483.

14. Bergstrom T, Bergstrom CT, Li H. Frequency and accuracy of proactive testing for COVID-19. medRxiv. 2020.

15. Wascher M, Schnell PM, Khudabukhsh WR, Quam M, Tien JH, Rempala GA. Monitoring SARS-COV-2 Transmission and Prevalence in Populations under Repeated Testing. medRxiv. 2021.

16. Rennert L, McMahan C, Kalbaugh CA, et al. Surveillance-based informative testing for detection and containment of SARS-CoV-2 outbreaks on a public university campus: an observational and modelling study. The Lancet Child & Adolescent Health. 2021;5(6):428–436.

17. Martin N, Schooley RT, De Gruttola V. Modelling testing frequencies required for early detection of a SARS-CoV-2 outbreak on a university campus. medRxiv. 2020.

18. Yamey G, Walensky RP. Covid-19: re-opening universities is high risk. British Medical Journal Publishing Group; 2020.

19. Wilson E, Donovan CV, Campbell M, et al. Multiple COVID-19 clusters on a university campus—North Carolina, August 2020. Morbidity and Mortality Weekly Report. 2020;69(39):1416.

20. Booeshaghi AS, Tan FH, Renton B, Berger Z, Pachter L. Markedly heterogeneous COVID-19 testing plans among US colleges and universities. medRxiv. 2020.

21. Lamont N. Executive Order No. 7C: Protection of Public Health and Safety During COVID-19 Pandemic and Response - Further Suspension or Modification of Statutes. State of Connecticut; 2020.

22. Ojakian M. Update #4 to the Higher Education Report: Recommendations for Reopening Undergraduate Colleges and Universities. Phase 3 Planning Framework for Reopenning for Reopening Undergraduate Residential Colleges and Universities: Higher Education Reopen Connecticut Subcommittee; 2020.

23. Hartocollis A, Hubler S. Covid Tests and Quarantines: Colleges Brace for an Uncertain Fall. New York Times; 2020.

24. Widness J. COVID-19 in Connecticut institutions of higher education. In: Crawford FW, ed 2021.

25. Connecticut Department of Public Health. Interim Guidance for the Reopening of Higher Education Campuses with On-Campus Residential Populations for the Spring 2021 Semester (01-08-2021). 2021.

26. Connecticut Department of Public Health. Interim Guidance for Surveillance Testing on Higher Education Campuses with On-Campus Residential Populations for the Spring 2021 Semester February 25, 2021. 2021.

27. Connecticut Department of Public Health. Interim Guidance for Surveillance Testing on Higher Education Campuses with On-Campus Residential Populations for the Spring 2021 Semester March 26, 2021. 2021.

28. Burt C. State-by-state look at colleges requiring COVID-19 vaccines. 2021; https://universitybusiness.com/state-by-state-look-at-colleges-requiring-vaccines/. Accessed July 12, 2021.

29. Student Health Services. COVID Vaccination Requirement for Residential Students and Student-Athletes University of Saint Joseph; 2021.

30. Leidner AJ, Barry V, Bowen VB, et al. Opening of large institutions of higher education and county-level COVID-19 incidence—United States, July 6–September 17, 2020. Morbidity and Mortality Weekly Report. 2021;70(1):14.

31. Lu H, Weintz C, Pace J, Indana D, Linka K, Kuhl E. Are college campuses superspreaders? A data-driven modeling study. Computer Methods in Biomechanics and Biomedical Engineering. 2020:1–11.

32. Ivory D, Smith M, Lee J, et al. See How Vaccinations Are Going in Your County and State. New York Times; 2021.

33. Fall 2020 Enrollment. Connecticut Office of Higher Education; 2020.

34. Widness J. Filling in CCIC Data. In: Schultes O, ed 2021.

35. Pritchard A. RE: follow-up on recent data request [not-secure]. In: Schultes O, ed 2021.

36. Shea W. FW: Data Request [not-secure]. In: Sosa L, ed 2021.

37. 2019 Connecticut Higher Education System Data and Trends Report. Hartford, CT: Connecticut Office of Higher Education; 2019.

38. Onorato S. Re: UConn COVID Testing Data. In: Schultes O, ed 2021.

39. COVID-19 Dashboard. 2021; https://coviddashboard.uconn.edu/covid-dashboard/#. Accessed July 12, 2021.

40. Crawford FW, Jones SA, Cartter M, et al. Impact of close interpersonal contact on COVID-19 incidence: evidence from one year of mobile device data. medRxiv. 2021:2021.2003.2010.21253282.

41. Morozova O, Li ZR, Crawford FW. One year of modeling and forecasting COVID-19 transmission to support policymakers in Connecticut. medRxiv. 2021:2020.2006.2012.20126391.

42. COVID-19: Appendices. 2021; https://www.cdc.gov/coronavirus/2019-ncov/php/contact-tracing/contact-tracing-plan/appendix.html#contact. Accessed July 15, 2021.

43. American Community Survey (ACS). 2021; https://www.census.gov/programs-surveys/acs. Accessed July 15, 2021.

44. Glossary. 2019; https://www.census.gov/programs-surveys/geography/about/glossary.html#par_textimage_4. Accessed July 15, 2021.

45. tidycensus: Load US Census Boundary and Attribute Data as ‘tidyverse’ and ‘sf’-Ready Data Frames [computer program]. Version R package version 0.11.42021.

46. United States Census Bureau. Ensuring an Accurate Count of College Students and Towns in the 2020 Census. 2020; https://www.census.gov/newsroom/press-releases/2020/2020-college-students.html. Accessed July 30, 2021.

47. R: A language and environment for statistical computing [computer program]. Version 1.2.1335. Vienna, Austria: R Foundation for Statistical Computing; 2019.

48. Lamont N. Executive Order No. 7III: Protection of Public Health and Safety During COVID-19 Pandemic and Response – Mandatory Self-Quarantine of Travelers Arriving from States with High COVID-19 Infection Rates and Extension of Certain Deadlines Applicable to the Department of Motor Vehicles. State of Connecticut; 2020.

49. Saul S. Indiana University Can Require Students to Get Coronavirus Vaccines. New York Times; 2021.

50. Brown CM. Outbreak of SARS-CoV-2 Infections, Including COVID-19 Vaccine Breakthrough Infections, Associated with Large Public Gatherings—Barnstable County, Massachusetts, July 2021. MMWR. Morbidity and Mortality Weekly Report. 2021;70.

51. Connecticut Department of Public Health. COVID-19_Tests__Cases__Hospitalizations__and_Deaths__Statewide_. 2021.

52. Currie DW, Moreno GK, Delahoy MJ, et al. Description of a University COVID-19 Outbreak and Interventions to Disrupt Transmission, Wisconsin, August-October 2020. medRxiv. 2021.

53. Andersen MS, Bento AI, Basu A, Marsicano CR, Simon K. College Openings, Mobility, and the Incidence of COVID-19. medRxiv. 2021.

54. Li Y, Ma C, Tang W, Zhang X, Zhu J, Nallamothu B. Association of university reopening policies with new confirmed covid-19 cases in the united states. medRxiv. 2021:2020.2012. 2011.20247353.

55. Richmond CS, Sabin AP, Jobe DA, Lovrich SD, Kenny PA. SARS-CoV-2 sequencing reveals rapid transmission from college student clusters resulting in morbidity and deaths in vulnerable populations. MedRxiv. 2020.

56. Guidance for Institutions of Higher Education (IHEs). 2021; https://www.cdc.gov/coronavirus/2019-ncov/community/colleges-universities/considerations.html. Accessed July 22, 2021.

57. Interim Guidance for SARS-CoV-2 Testing and Screening at Institutions of Higher Education (IHEs). 2021; https://www.cdc.gov/coronavirus/2019-ncov/community/colleges-universities/ihe-testing.html#ref4. Accessed July 22, 2021.

58. American College Health Association. Considerations for Reopening Institutions of Higher Education for the Fall Semester 2021. Silver Spring, MD: ACHA; 2021:1-34.

59. Johnson P, Brown R, Bunis D, et al. Report and Recommendations of the Massachusetts Higher Education Testing Group. MA Higher Education Testing Group; 2021.

60. Putterman A, Brindley E. Gov. Ned Lamont extends COVID-19 state of emergency in Connecticut; State’s testing numbers on the rise. Hartford Courant; 2020.

61. Betancourt WQ, Schmitz BW, Innes GK, et al. COVID-19 containment on a college campus via wastewater-based epidemiology, targeted clinical testing and an intervention. Science of The Total Environment. 2021;779:146408.

62. Karthikeyan S, Nguyen A, McDonald D, et al. Rapid, large-scale wastewater surveillance and automated reporting system enabled early detection of nearly 85% of COVID-19 cases on a University campus. medRxiv. 2021.

63. Gstalder S. ALBERTUS MAGNUS COLLEGE COVID-19 SPRING 2021 PLANS. Albertus Magnus College; 2021.

64. Barr S. In Historic 95th Anniversary Year, Vigilance Pays Off As Albertus Magnus College Reaches Thanksgiving Break. Albertus Magnus College; 2020.

65. Heitz ET, Magenheimer E. Albertus Magnus College; 2020.

66. Cintorino S. CCSU Blueprint: Plans for a Successful Fall 2020 Reopening. Central Connecticut State University; 2020.

67. Cintorino S. CCSU Blueprint: Plans for a Successful Spring 2021 Reopening. Central Connecticut State University; 2020.

68. Arcelus V, McKnight J, Singer J. Important Conn Fall 2020 Information (pre-arrival requirements, campus testing andmove-in). Connecticut College; 2020.

69. Arcelus V, McKnight J, Singer J. Important Information for Spring 2021 Return. Connecticut College; 2020.

70. Campus Life in Spring 2021. 2021; https://www.conncoll.edu/campus-life/student-health-services/coronavirus/campus-life-with-covid-19/. Accessed May 17, 2021.

71. COVID-19 Response Team. Returning to Conn, Frequently Asked Questions. 2021; https://www.conncoll.edu/campus-life/student-health-services/coronavirus/returning-to-conn-faqs/. Accessed July 23, 2021.

72. DeLisa K, Rose-Zak S. Eastern Connecticut State University Reopening Plan, Fall 2020. Eastern Connecticut State University; 2020.

73. DeLisa K, Rose-Zak S. Eastern Connecticut State University Reopening Plan, Spring 2021. Eastern Connecticut State University; 2020.

74. Housing and Student Life. 2021; https://www.easternct.edu/reopening/student-support/housing.html. Accessed July 22, 2021.

75. Lawlor K. Reopening Plans for Undergraduate Residential Colleges and Universities in Phase 3 - Name of Institution: Fairfield University. Fairfield University; 2020.

76. Plans for Fairfield’s Spring 2021 Return to Campus. 2020; https://www.fairfield.edu/news/archive/2021/january/spring-reopening-return-to-campus.html?utm_source=watson&utm_medium=social&utm_campaign=mc&utm_term=spring-2021&utm_content=fairfield-university-reopening. Accessed June 11, 2021.

77. Donoghue K. A Message from the Vice President for Student Life. Fairfield University; 2021.

78. Wright C. Mitchell_College_State_plan_Phase3.pdf. Mitchell College; 2020.

79. Spring MiniMester Courses: May + June. 2021; https://mitchell.edu/wp-content/uploads/2021/05/May-June-MiniMester-2021-to-REGISTERED-CURRENT-students.pdf. Accessed June 14, 2021, 2021.

80. Information about Returning to Campus this Spring! : Mitchell College; 2020.

81. Zemba B. Quinnipiac University Reopening Plan Phase 3 Fall 2020. Quinnipiac University; 2020.

82. Drucker M. Return to Campus Guide for Students Spring 2021. Quinnipiac University; 2021.

83. 8 new things you need to know for the spring. 2021; https://www.qu.edu/quinnipiac-today/8-new-things-you-need-to-know-for-the-spring-2021-01-12/. Accessed July 22, 2021.

84. MacNamara G. Sacred Heart University Undergraduate Residential Phase 3 Reopening Campus Plan Fall 2020.Sacred Heart University; 2020.

85. Community Updates. Sacred Heart University; 2021.

86. Richardson A. Reopening Plans for Colleges and Universities in Phase 3 July 28, 2020. Southern Connecticut State University; 2020.

87. Richardson A. Reopening Plans for Colleges and Universities in Phase 3 January 2021. Southern Connecticut State University; 2021.

88. Berger-Sweeney J, Rojas J. Trinity College Health and Safety Plan for Reopening. Trinity College; 2020.

89. DiChristina J, Goodman J. Update on PCR COVID 19 Testing and State of Connecticut Travel Advisory. Trinity College; 2020.

90. DiChristina J, Rojas J. Preparing for the spring semester at Trinity College. Trinity College; 2021.

91. CoVerified App - Reopening. 2021; https://www.trincoll.edu/reopening/coverified-app/. Accessed June 14, 2021.

92. Schmidt R. University of Bridgeport Re-opening Plan Phase 3. University of Bridgeport; 2020.

93. Frequently Asked Questions. 2021; https://www.bridgeport.edu/covid-19/faq. Accessed June 14, 2021.

94. Sanders C. Important Move-In Information. University of Bridgeport; 2021.

95. Jordan S. Reopen Phase 3 Fall Semester Commencing on or after 14 August 2020. University of Connecticut; 2020.

96. Jordan S. Reopen Plan Spring Semester Commencing 16 January 2021. University of Connecticut; 2021.

97. Nicklin J. Reopening Plan for the University of Hartford in Phase 3 - Fall 2020. University of Hartford; 2020.

98. Testing for COVID-19. 2021; https://www.hartford.edu/healthy-hawks/health-safety-guidelines/covid-testing.aspx#accordion-group-1-section-1-label. Accessed June 11, 2021.

99. Office of Marketing and Communication. Move-In Updates. University of Hartford; 2021.

100. McGee S. Reopening Plan - Fall 2020. University of New Haven; 2020.

101. McGee S. University Response to COVID-19: Important Notifications and FAQ. 2021; https://www.newhaven.edu/covid19/notifications-and-faq.php. Accessed June 11, 2021.

102. McGee S. The Phased Plan for Return to Campus - Spring 2021: Phase 3. University of New Haven.

103. Free R. Reopening Plans for Colleges and Universities in Phase 3 - Fall 2020. University of Saint Joseph; 2020.

104. DiSalvo E. Colleges Start Spring Semester with Souped up Testing. 2021; https://we-ha.com/colleges-start-spring-semester-with-souped-up-testing/. Accessed July 23, 2021.

105. Culliton R. Reopening Plans for Undergraduate Residential Colleges and Universities in Phase 3. Wesleyan University; 2020.

106. All About Testing. Wesleyan University.

107. Whaley M, Roth M. Important Information for Fall Semester. Wesleyan University; 2020.

108. Safety Guidelines. Wesleyan University.

109. Prepare for Arrival. 2021; https://www.wesleyan.edu/reactivatingcampus/prepare-arrival.html. Accessed July 22, 2021.

110. Koukopoulos P. Reopening Plans for Colleges and Universities in Phase 3 - Fall 2020. Western Connecticut State University; 2020.

111. Koukopoulos P. Reopening Plans for Colleges and Universities in Phase 3 - Spring 2021. Western Connecticut State University; 2020.

112. Spangler S, Loucks N. Yale University “Reopen Connecticut” Phase 3 Plan. Yale University; 2020.

113. Boyd M. Spring 2021 housing and move-in information (December 8). Yale University; 2020.

114. Chen C. Students: Pre-Arrival Testing for Spring 2021. Yale University; 2021.

115. Yale Community Compact. Yale University; 2021.

